# Vaccination against COVID-19 and society’s return to normality in England: a modelling study of impacts of different types of naturally acquired and vaccine induced immunity

**DOI:** 10.1101/2021.05.18.21257314

**Authors:** Fujian Song, Max O. Bachmann

## Abstract

**Objectives:** To project impacts of mass vaccination against COVID-19, and investigate possible impacts of different types of naturally acquired and vaccine-induced immunity on future dynamics of SARS-CoV-2 transmission from 2021 to 2029 in England.

**Design:** Deterministic, discrete-time population dynamic modelling.

**Participants:** Population in England.

**Interventions:** mass vaccination programmes.

**Outcome measures:** daily and cumulative number of deaths from COVID-19.

**Results:** If vaccine efficacy is ≥ 70%, the vaccine-induced sterilising immunity lasts ≥ 182 days, and the reinfectivity is greatly reduced (by ≥ 40%), mass vaccination programmes can prevent further COVID-19 outbreaks in England. Under such optimistic scenarios, the cumulative number of COVID-19 deaths is estimated to be from 113,000 to 115,000 by the end of 2029 in England. However, under plausible scenarios with lower vaccine efficacy, shorter durability of immunity, and smaller reduction in reinfectivity, repeated vaccination programmes could not prevent further COVID-19 outbreaks.

**Conclusions:** Under optimistic scenarios, mass immunisation using efficacious vaccines may enable society safely to return to normality. However, under plausible scenarios with low vaccine efficacy and short durability of immunity, COVID-19 could continue to cause recurrent waves of severe morbidity and mortality despite frequent vaccinations. It is crucial to monitor the vaccination effects in the real world, and to better understand characteristics of naturally acquired and vaccine induced immunity against SARS-CoV-2.

**ARTICLE SUMMARY:** *Strengths and limitations of this study:*

- We used a population dynamic model to assess impacts of vaccination programmes on future dynamics of SARS-CoV-2 transmission dynamics, and to explicitly investigate the impacts of different types of immune responses to SARS-CoV-2 infection and vaccines on the COVID-19 epidemic in England.
- The model has been verified based on historically observed outcome data in England, and a large number of projection scenarios are explored.
- Findings from our study improves the understanding of key immunological parameters relevant to future SARS-CoV-2 transmission dynamics and vaccination strategies.
- This is a deterministic simulation model, and uncertainty in estimated parameters may have not been fully accommodated. There remain many uncertainties regarding durability and types of naturally acquired and vaccine-induced immunity.

## INTRODUCTION

The COVID-19 pandemic caused by the spread of SARD-CoV-2 virus has resulted in a huge number of deaths and severe disruptions of economies and social activities around the world. The spread of the SARS-CoV-2 virus can be suppressed by non-pharmaceutical interventions (NPIs) and lockdown measures.^1^ Because of their disruptive socioeconomic consequences, lockdown restrictions cannot last indefinitely.

Only a few months after the initial identification of SARS-CoV-2 pathogen, there were more than 200 vaccine candidates in development globally.^2^ Since December 2020, three vaccines against COVID-19 have been approved for use in the UK, and a vaccination programme has been started to rollout, prioritised primarily by age and comorbidity, with older people being vaccinated first.^3^ Although mass vaccination is a promising strategy to enable society to safely return to normality, there is great uncertainty about COVID-19 vaccines, including their safety and efficacy, and durability of different types of protection after vaccination.

The protection of naturally acquired or vaccine-induced immune responses may be attributable to infection protection, disease reduction, and reinfectivity reduction.^4^ Studies of diseases caused by other human coronaviruses (HCoVs) indicated that infection protection immunity is likely to be short-lived, while disease reduction and reinfectivity reduction are likely long lasting.^5^ Therefore, we conducted a modelling study to investigate possible impacts of different types of naturally acquired and vaccine-induced immunity on future dynamics of SARS-CoV-2 transmission in England.

## METHODS

### Model structure

This is a deterministic discrete-time (day) population dynamic model, implemented with computational language R.^6^ The population are classified into categories by sex, age (5-year age bands for age <10 years, and 10-year age bands for age ≥ 10 years), and COVID-19 infection status (figure 1). The main infection compartments include susceptible, exposed, infectious, recovered, and vaccinated. Here “exposed” refers to a pre-infectious status of infected individuals. Infected individuals are classified as asymptomatic or symptomatic, and symptomatic individuals are classified as not being isolated, self-isolated, and hospitalised. We assume that hospitalised patients are effectively isolated and no longer able to transmit the virus to the general population, but patients who self-isolate at home may transmit virus to household contacts. The recovered and vaccinated are protected from reinfection, but they may be reinfected if the immunity is short-lived.

**Figure 1:**
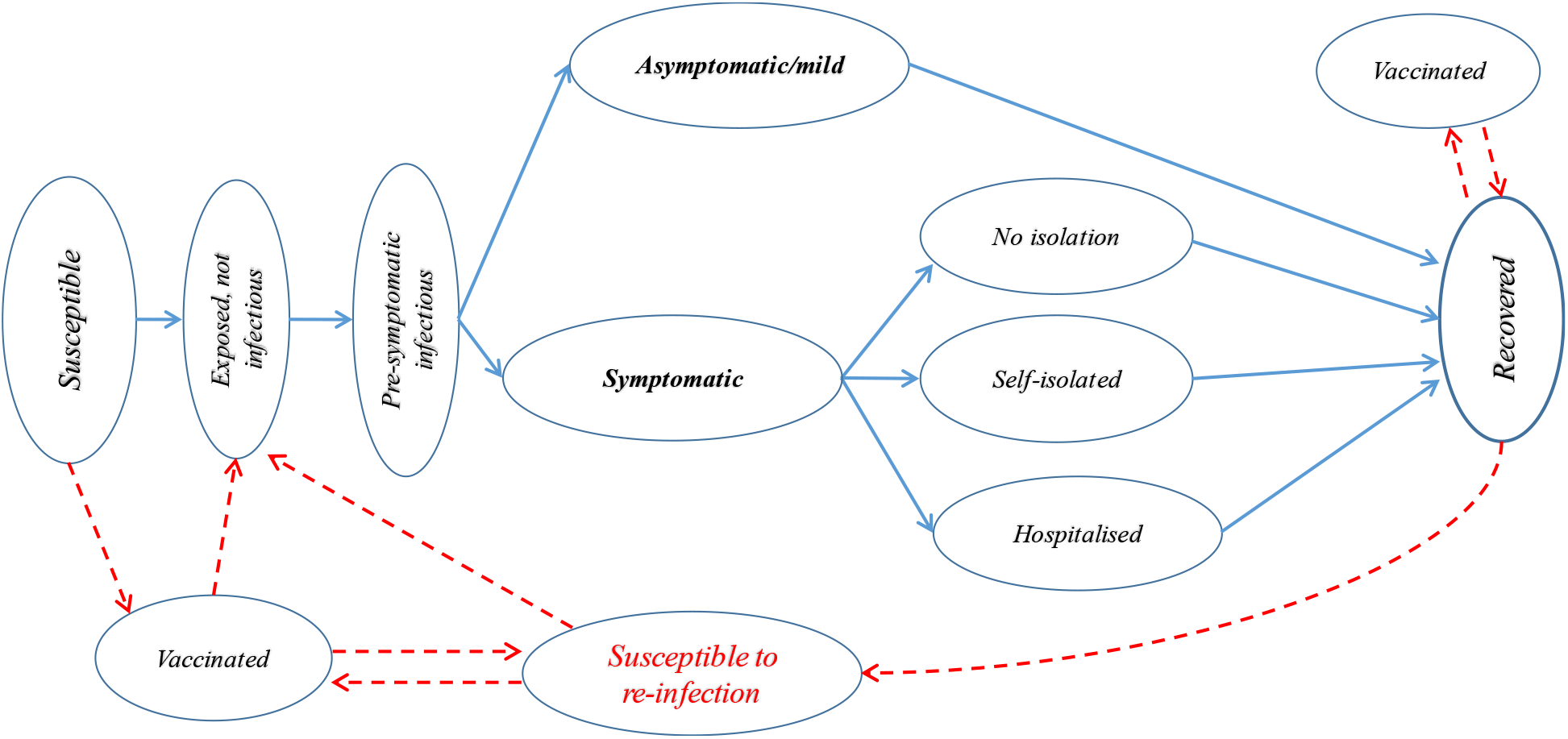
Modelling COVID-19 epidemics in England - main compartments and transitions across status.

### Parameters and data sources

Details on the model’s structure (appendix figure 1), parameters (appendix table 1), source of data, and mathematical equations are available in Supplementary files. Initial parameter values were estimated based on a review of relevant literature, and key parameter estimates were adjusted so that the simulated numbers of COVID-19 deaths, hospitalised patients, and recovered individuals were as similar as possible to the data historically reported from March 2020 to January 2021 in England.^7^

We obtained population demographic statistics in England from the Office for National Statistics,^8^ and the whole population is assumed to be susceptible to SARS-CoV-2 infection at the beginning of 2020. We assume incubation periods, infectious periods, days of hospital stay, and days of deaths after being infected, to be gamma distributed.^9-11^ Age specific case fatality rates and hospitalisation rates of symptomatic cases were based on a study by Verity et al.^11^ Average sex-and-age-specific rates of all-cause deaths in England during 2015-2019^8^ are applied to people who are not infected with or recovered from COVID-19. We assumed that the number of births equals to the number of deaths each day, and did not consider the influence of population migration. We adjusted the number of individuals belonging to each age group at the beginning of a year since 2021, by shifting a proportion of them to the adjacent higher age group.

For simplicity, effects of NPI measures, including restrictions on social activities, contact tracing and testing, were materialised by changes in transmission risk per contact between a susceptible and an infectious individual,^12^ and average numbers of contacts of the general population (appendix table 2 in supplementary files). We estimate that the transmission risk per contact between infectious and susceptible individuals was reduced by 32%, from 0·068 before the implementation of any NPIs to 0·046 by March 15, 2020 after implementing basic NPI measures. Because of the new virus variant (B.1.1.7),^13^ the average transmission risk per contact was increased to 0·056 by the end of 2020. We assume that the transmission risk per contact from April to September is 20% lower than that from October to February, to incorporate the impact of seasonality on future projections since April 2021. We do not use the reproduction number as an input parameter, but derived the basic and effective reproduction numbers based on model’s transmission parameters (equation 79 in Supplementary files).^12 14^

The sex-and-age-specific numbers of daily contacts per person were based on the UK data from a study of European countries.^15^ We consider only the daily contacts of the general population and household contacts of individuals self-isolated at home. We estimated that the lockdown measures from March 24, 2020 reduced general population contacts by 60-85%, although household contacts were unchanged. The NPI measures were relaxed or strengthened over time, which were reflected in the assumed social contacts and transmission risk (Supplementary files). After rolling out vaccination, we assume that, since June 2021, social contracts are return to normal as before the pandemic in England, although basic hygienic measures would be maintained.

### Vaccination and projection scenarios

Results of randomised controlled trials shown that vaccines were efficacious in reducing symptomatic diseases, compared with placebo.^16 17^ Vaccination of prioritised individuals began from 8 December 2020 in England and around 2 million individuals were vaccinated (mostly with a single dose) by January 10, 2021.^18^ For simplicity, we assume that the mass vaccination starts from January 1, 2021 with a 80% coverage of eligible individuals, and the maximum number of individuals vaccinated daily is 300,000. The mass vaccination is modelled as an age-based phase approach, starting from people aged ≥ 70, followed by individuals aged 60-69, 50-59, 20-49, and then those aged 16-19. Although both Pfizer-BioNTec and AstraZeneca vaccines are 2-dose regimens, the policy in the UK has been to initially provide the first dose to as many adults as possible. Data from clinical trials indicated that the short-term vaccine efficacy after the first dose of the Pfizer-BioNTech and the AstraZeneca vaccine is, respectively, about 90% and 70%.^16^ For simplicity, we do not separate single or double dose vaccination, and assume a range of the overall vaccine efficacy (90%, 70% or 50%) and the protection effects start 14 days after vaccination.

The reduction in symptomatic cases in vaccinated individuals may be due to induced antibodies in susceptible individuals (infection protection), or a lower proportion of infected individuals being symptomatic (i.e., disease reduction), or a combination of both. There are many different possible combinations of infection protection and disease reduction for a given overall vaccine efficacy in reducing symptomatic cases (appendix figure 2 in supplementary files). We assume that vaccine efficacy for reducing symptomatic cases is equally attributable to infection reduction and disease reduction in the main projections. For vaccines with 90%, 70% and 50% overall efficacy, the equal partial efficacy for the infection protection and for disease reduction is around 69%, 45%, and 29%, respectively (see Supplementary files for details).

Immune responses against COVID-19 infection, either naturally acquired from past infection or vaccine-induced, may reduce individuals’ susceptibility to infection (sterilising or infection protection immunity), reduce pathology so that disease is less severe after being infected (disease reduction immunity), and reduce infectivity of those who are reinfected after the waning of immunity (reinfectivity reduction immunity).^4^ According to immunological characteristics of other HCoVs, infection protection immunity may wane after a short period, while disease protection and reinfectivity reduction immunity are likely longer lasting.^5^ For example, antibodies against SARS-CoV-1 virus in recovered patients was no longer detectable after 2-3 years, while specific memory T cells remained detected after 11 years.^19^ Therefore, we assume that the disease reduction and reinfectivity reduction immunity are long lasting (>10 years).^5^ We assume that naturally acquired sterilising immunity lasts for 365 or 730 days, and vaccine-induced sterilising immunity lasts for 182, 365 or 730 days. Available evidence indicated that the viral loads and the duration of virus shedding in the infected individuals after vaccination were considerably reduced, compared with unvaccinated individuals.^20 21^ Therefore, we assume that reinfectivity after waning of sterilising immunity is reduced by 20%, 40% or 60%. In this study, we assume that the infectivity of ineffectively vaccinated individuals is the same as individuals after waning of sterilising immunity, and vaccination of individuals recovered from natural infection prevents or delays the waning of their sterilising immunity.

We run the model and calibrate key transmission parameters by visually comparing estimated numbers of daily COVID-19 deaths, and hospitalised patients, with official records from January 1, 2020 to January 31, 2021 in England. We used estimates of transmission parameters by the end of January 2021 to project COVID-19 deaths from 2021 to 2029, under various scenarios of vaccine efficacy, durability and protection characteristics of naturally acquired and vaccine-induced immunity. The number of deaths from SARS-CoV-2 infections is the main endpoint in this study.

### Patient and public involvement

No patients and the public were involved in this literature and secondary data based, computational modelling study.

## RESULTS

Our derived basic reproduction number (R0) was 3·68 at the initial stage of the COVID-19 epidemic in England (appendix figure 3 in supplementary files). After implementing NPI and lockdown measures, the effective reproduction value (Rt) was reduced to 0·66 from March 24, 2020. Thereafter, the Rt values fluctuated along with changing NPI policies, and our estimated R values during March 2020 and January 2021 were within the ranges reported in England (Appendix figure 3 in Supplementary files).^22^ The estimated prevalence of SARS-CoV-2 infection (appendix figure 4), the number of hospitalised COVID-19 patients (appendix figure 5), and the estimated daily deaths from COVID-19 (appendix figure 6) are well matched with the reported data from March 2020 to January 2021 in England (Supplementary files).

### Vaccine efficacy, immunity durability, and reinfectivity

Figure 2 shows the impacts of partial vaccine efficacy regarding disease reduction relative to infection protection, durability of immunity, and reinfectivity, given the same overall vaccine efficacy (70%) in reducing symptomatic cases. There are three general inferences. As expected, the number of COVID-19 deaths is smaller following a greater reduction in reinfectivity (figure 2A, 2B, 2C). Second, a greater reduction in reinfectivity makes the durability of immunity less influential, if a vaccine is efficacious for infection protection (figure 2c). Third, the greater the infection protection by a vaccine, the smaller the number of COVID-19 deaths, although there are exceptions (figure 2A). A combination of a shorter duration of immunity and smaller reduction in the reinfectivity makes the disease reduction efficacy more important.

**Figure 2.**
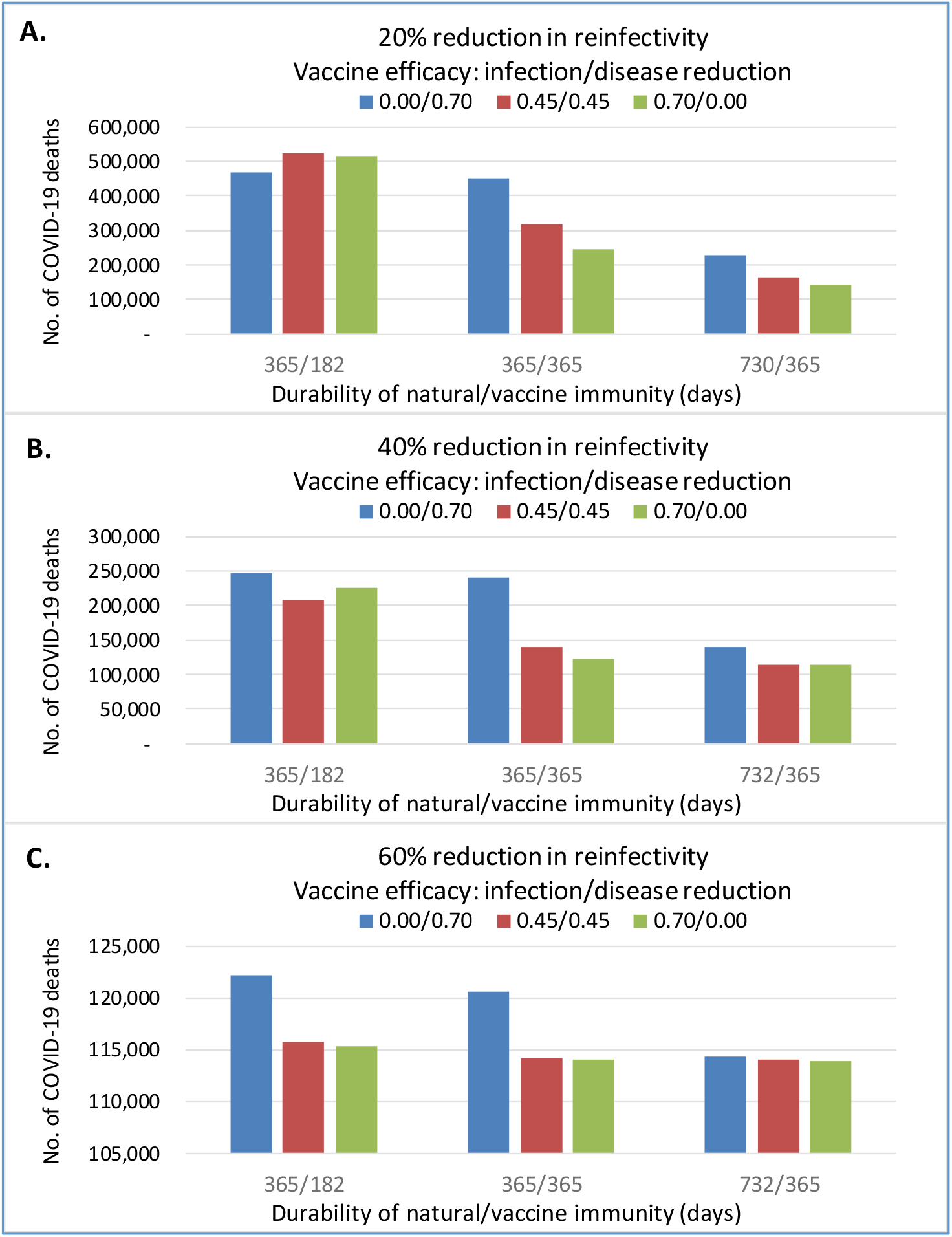
Projected numbers of COVID-19 deaths with different combinations of infection and disease protection vaccine efficacy, durability of immune response, and reduction in reinfectivity. The cumulative number of COVID-19 deaths in England by the end of 2029, after nine repeated annual vaccinations of 80% individuals aged ≥ 16 during 2021-2029. The overall vaccine efficacy was 70%; “0·00/0·70” refers to all vaccine efficacy attributable to disease protection, “0·45/0·45” refers to equal infection and disease protection, “0·70/0·00” refers to all efficacy attributable to infection protection. Duration of immunity: “365/182” refers to 365 days of natural immunity and 182 days vaccine immunity, “365/365” refers to 365 days natural and vaccine immunity, “730/365” refers to 730 and 365 days, respectively, natural and vaccine immunity. Figure 2A, 2B and 2C shows results under the assumption of 20%, 40% and 60% reduction in reinfectivity.

### Population susceptibility and COVID-19 outbreaks

Changes in the prevalence of susceptible individuals and daily peaks of COVID-19 deaths in England during 2020 and 2029 are shown in figure 3 (additional details in supplementary table 1), under assumptions of 90% vaccine efficacy, 40% reduction in reinfectivity after waning of sterilising immunity, and different durations of naturally acquired and vaccine-induced sterilising immunity.

**Figure 3.**
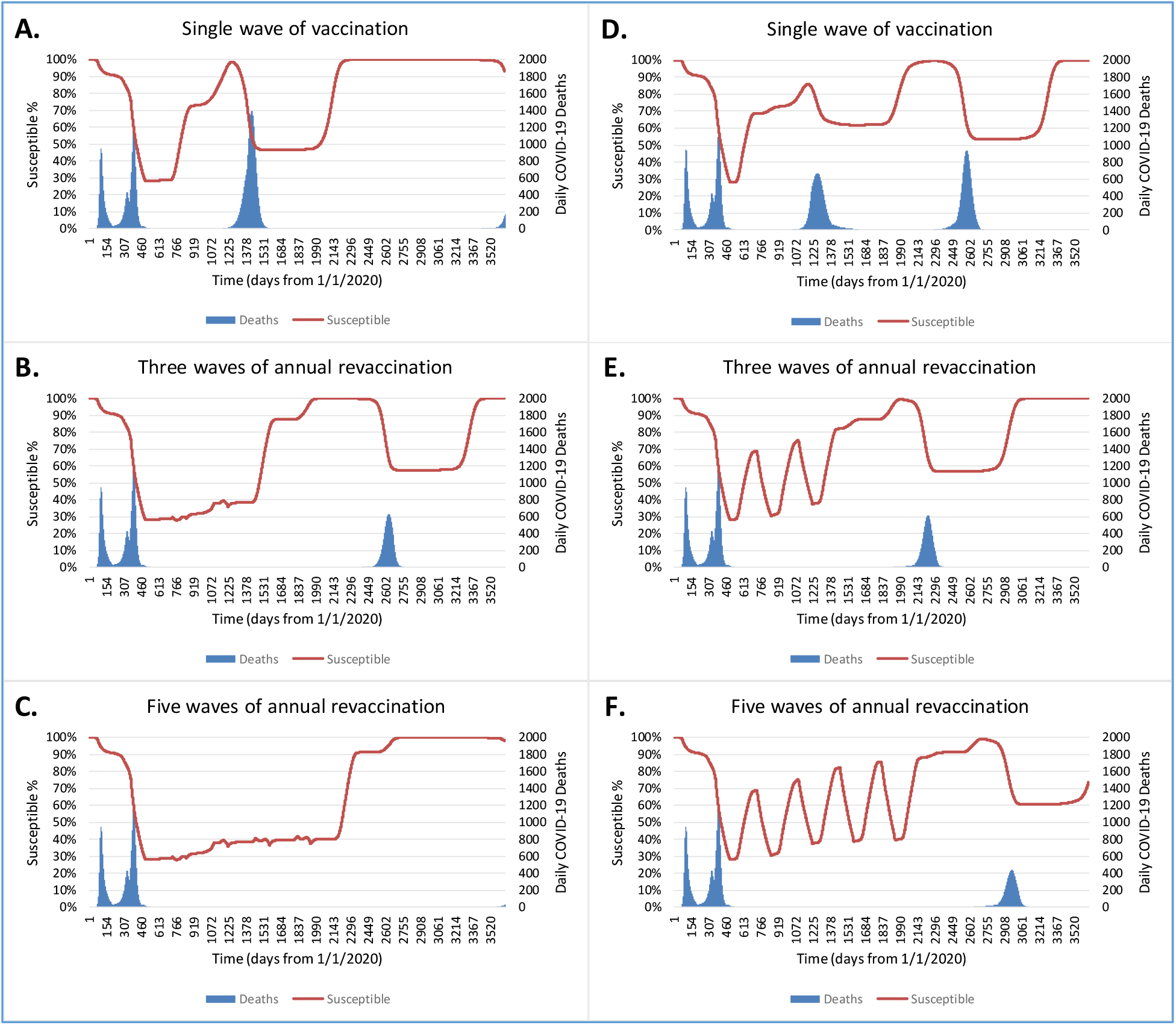
Projected peaks of daily COVID-19 deaths and the prevalence of susceptible individuals (%) during 2020-2029 under scenarios with different immunity durability and vaccine strategies. We assume 90% vaccine efficacy; 80% coverage of individuals aged ≥ 16; 40% reduction in reinfectivity and 730 days natural immunity for all scenarios in Figure3. We assume 365 days (Figure 3A, 3B, 3C) or 182 days (Figure 3D, 3E, 3F) vaccine immunity, and single vaccination (Figure 3A, 3D), three annual vaccination (Figure 3B, 3E), or five annual vaccination programmes (Figure 3C, 3F).

The prevalence of susceptible individuals is reduced to <30% after a single wave of mass vaccination starting from January 2021, but starts to increase from January 2022, if the duration of vaccine immunity is 365 days (figure 3A). The raised prevalence of susceptible individuals leads to an outbreak with a high peak (n=1,394) of daily COVID-19 deaths in November 2023. The prevalence of the susceptible is reduced to about 47% by the natural infection during the outbreak, then increases to >90% by the end of 2025 due to waning of immunity, which is followed with a new outbreak from December 2029. Clearly, a single wave of mass vaccination is insufficient if the immune response is short-lived. Three waves of repeated annual revaccination programmes delay the new outbreak, with a peak (n=632) of daily COVID-19 deaths in March 2027 (figure 3B). Five repeated waves of annual revaccination programmes almost prevent any new outbreaks before the end of 2029 (figure 3C).

If the vaccine immunity lasts only 182 days, the prevalence of susceptible individuals starts to increase six months after vaccination (figure 3D, 3E, 3F). A single wave of mass vaccination is followed with two new peaks of daily COVID-19 deaths (figure 3D), and the three and five repeated waves of annual vaccination programmes result in corresponding changes in the prevalence of susceptible individuals, each with a single peak of daily COVID-19 deaths (figure 3E, 3F). If the vaccine immunity lasts only 182 days, the annual mass vaccination programmes are insufficient to sustain a constantly low prevalence of the susceptible, and the prevalence of the susceptible fluctuates up and down biannually (figure 3E, 3F). Notably, the COVID-19 outbreaks start about two years after stopping the annual mass vaccination programmes, due to the assumed duration of natural immunity (730 days).

### Total COVID-19 deaths under various scenarios

The projected total numbers of COVID-19 deaths during 2020-2029, under various scenarios, are shown in table 1 (more details in Supplementary table 2). If there no waning of immunity, a single mass vaccination programme prevents COVID-19 outbreaks after returning to normality. If there is waning of sterilising immunity, mass vaccination programmes may prevent further COVID-19 outbreaks under scenarios with high vaccine efficacy, longer lasting immunity, and large reduction in reinfectivity after waning of sterilising immunity. Under optimistic scenarios, the cumulative number of COVID-19 deaths is estimated to be from 113,000 to 115,000 by the end of 2029 in England. However, the total number of COVID-19 deaths may be up to 754,000 by the end of 2029, under the scenario with low vaccine efficacy (50%), short duration of sterilising immunity, and high reinfectivity (table 1).

**Table 1.** Projected total COVID-19 deaths by the end of 2029 in England under various scenarios. Notes to Table 1: We assume annual vaccination covers 80% of individuals aged ≥ 16 years, and the overall vaccine efficacy is equally attributable to the infection and disease protection. The maximum number of waves of annual mass vaccination is nine during 2021-2029. Results in the table are from scenarios with the possibly minimum numbers of waves of annual vaccination that prevent any further COVID-19 outbreaks during 2022-2029 under a given scenario, or those in which further COVID-19 outbreaks cannot be prevented by the maximum number of annual mass vaccination programmes.

| Scenario | Waves of annual vaccination | Overall vaccine efficacy | Reduction in reinfectivity | Immunity duration (days) |  | Total COVID-19 deaths (2020-2029) |
| --- | --- | --- | --- | --- | --- | --- |
|  |  |  |  | Natural | Vaccine |  |
| st1-01 | 1 | 90% | 60% | >10 years | >10 years | 112,815 |
| st1-02 | 2 | 90% | 60% | 730 | 730 | 112,815 |
| st1-03 | 2 | 90% | 60% | 730 | 365 | 112,815 |
| st1-04 | 2 | 90% | 60% | 730 | 182 | 112,815 |
| st1-05 | 3 | 90% | 60% | 365 | 365 | 112,870 |
| st1-06 | 3 | 90% | 60% | 365 | 182 | 112,870 |
| st1-07 | 1 | 90% | 40% | >10 years | >10 years | 112,878 |
| st1-08 | 5 | 90% | 40% | 730 | 730 | 112,878 |
| st1-09 | 6 | 90% | 40% | 730 | 365 | 112,878 |
| st1-10 | 9 | 90% | 40% | 730 | 182 | 112,878 |
| st1-11 | 7 | 90% | 40% | 365 | 365 | 112,986 |
| st1-12 | 9 | 90% | 40% | 365 | 182 | 112,986 |
| st1-13 | 1 | 90% | 20% | >10 years | >10 years | 112,942 |
| st1-14 | 7 | 90% | 20% | 730 | 730 | 112,942 |
| st1-15 | 9 | 90% | 20% | 730 | 365 | 114,351 |
| st1-16 | 9 | 90% | 20% | 730 | 182 | 137,484 |
| st1-17 | 9 | 90% | 20% | 365 | 365 | 133,713 |
| st1-18 | 9 | 90% | 20% | 365 | 182 | 237,701 |
| st1-19 | 1 | 70% | 60% | >10 years | >10 years | 114,065 |
| st1-20 | 2 | 70% | 60% | 730 | 730 | 114,065 |
| st1-21 | 3 | 70% | 60% | 730 | 365 | 114,065 |
| st1-22 | 3 | 70% | 60% | 730 | 182 | 114,065 |
| st1-23 | 3 | 70% | 60% | 365 | 365 | 114,223 |
| st1-24 | 5 | 70% | 60% | 365 | 182 | 115,782 |
| st1-25 | 1 | 70% | 40% | >10 years | >10 years | 114,271 |
| st1-26 | 6 | 70% | 40% | 730 | 730 | 114,271 |
| st1-27 | 8 | 70% | 40% | 730 | 365 | 114,271 |
| st1-28 | 9 | 70% | 40% | 730 | 182 | 129,197 |
| st1-29 | 9 | 70% | 40% | 365 | 365 | 139,064 |
| st1-30 | 9 | 70% | 40% | 365 | 182 | 208,516 |
| st1-31 | 1 | 70% | 20% | >10 years | >10 years | 115,004 |
| st1-32 | 9 | 70% | 20% | 730 | 730 | 128,318 |
| st1-33 | 9 | 70% | 20% | 730 | 365 | 163,962 |
| st1-34 | 9 | 70% | 20% | 730 | 182 | 200,252 |
| st1-35 | 9 | 70% | 20% | 365 | 365 | 316,166 |
| st1-36 | 9 | 70% | 20% | 365 | 182 | 524,396 |
| st1-37 | 1 | 50% | 60% | >10 years | >10 years | 115,414 |
| st1-38 | 3 | 50% | 60% | 730 | 730 | 115,414 |
| st1-39 | 3 | 50% | 60% | 730 | 365 | 115,414 |
| st1-40 | 3 | 50% | 60% | 730 | 182 | 115,419 |
| st1-41 | 9 | 50% | 60% | 365 | 365 | 117,163 |
| st1-42 | 9 | 50% | 60% | 365 | 182 | 125,400 |
| st1-43 | 1 | 50% | 40% | >10 years | >10 years | 116,605 |
| st1-44 | 9 | 50% | 40% | 730 | 730 | 116,693 |
| st1-45 | 9 | 50% | 40% | 730 | 365 | 135,263 |
| st1-46 | 9 | 50% | 40% | 730 | 182 | 165,916 |
| st1-47 | 9 | 50% | 40% | 365 | 365 | 236,326 |
| st1-48 | 9 | 50% | 40% | 365 | 182 | 316,759 |
| st1-49 | 1 | 50% | 20% | >10 years | >10 years | 139,385 |
| st1-50 | 9 | 50% | 20% | 730 | 730 | 213,589 |
| st1-51 | 9 | 50% | 20% | 730 | 365 | 239,474 |
| st1-52 | 9 | 50% | 20% | 730 | 182 | 275,955 |
| st1-53 | 9 | 50% | 20% | 365 | 365 | 578,710 |
| st1-54 | 9 | 50% | 20% | 365 | 182 | 754,019 |

If the overall vaccine efficacy is 90%, the natural immunity lasts 730 days on average and the reinfectivity after immunity waning is reduced by 60%, two repeated annual vaccination programmes are able to prevent further outbreaks, even if the duration of the vaccine immunity is only 180 days. However, if the reinfectivity is reduced by 40%, five to nine annual revaccination programmes are required, depending on durability of immunity. If both the natural and vaccine immunity are short-lived (365 day and 180 days, respectively), and the reduction in reinfectivity is smaller (e.g., by 40% or 20%), further COVID-19 outbreaks cannot be prevented by repeated annual vaccination programmes (table 1).

If vaccine efficacy is 70%, the duration of natural immunity lasts 730 days and the reinfectivity is reduced by 60%, two to three annual vaccination programmes can prevent further COVID-19 outbreaks (table 1). If the sterilising immunity is short-lived (for 365 or 182 days only) and the reduction in reinfectivity is low (e.g., by only 20%), the annual mass vaccination programmes are not able to prevent further COVID-19 outbreaks. Similarly, when the vaccine efficacy is 50%, annual mass vaccination programmes prevent further COVID-19 outbreaks, only if the sterilising immunity is long lasting and the reduction in reinfectivity is large.

### Intervals between immunisation programmes

Results of sensitivity analyses based on vaccination programmes every two years or twice a year are shown in supplementary table 3. Programmes every two years can prevent COVID-19 outbreaks in scenarios with longer duration of sterilising immunity and larger reduction in reinfectivity. In addition, biannual vaccination programmes (twice a year) are more effective in reducing COVID-19 deaths, compared with annual programmes, in scenarios with low vaccine efficacy, short duration of sterilising immunity, and small reduction in reinfectivity. For example, in the scenario of 70% vaccine efficacy, 365 and 182 days of natural and vaccine immunity respectively, and 20% reduction in reinfectivity, the total number of COVID-19 deaths is estimated to be around 524,000 by annual vaccination programmes (table 1), which is reduced to around 228,000 by biannual vaccination programmes (Supplementary table-3). However, because a wave of mass vaccination of 80% individuals aged ≥ 16 years takes more than five months to complete, repeated vaccination every six months may not be practically feasible.

## DISCUSSIONS

Mass immunisation using efficacious vaccines may enable society safely to return to normality. Repeated vaccination programmes may be required to prevent further COVID-19 outbreaks, depending on vaccine efficacy, the durability and characteristics of different types of immune response to naturally acquired and vaccine-induced immune responses. Evidence on diseases caused by other common HCoVs indicated that the infection protection immunity may be short-lived, but the disease reduction and the reinfectivity reduction immunity are likely to be longer lasting.^5^ We found that, if reinfectivity is greatly reduced (e.g., by 60%), two or three repeated annual mass vaccination programmes prevent further COVID-19 outbreaks, even if the vaccine induced sterilising immunity lasts only 182 days. Under such optimistic scenarios, the cumulative number of COVID-19 deaths during 2020-2029 in England is estimated to be around 113,000. If both the natural and vaccine immunity are short-lived (365 and 180 days, respectively), and reinfectivity is reduced only by 40% or 20%, further COVID-19 outbreaks cannot be prevented by annual vaccination programmes. The total number of COVID-19 deaths is estimated to be around 754,000 by the end of 2029, under a pessimistic scenario with low vaccine efficacy (50%), short duration of sterilising immunity, and high reinfectivity.

There were several published modelling studies of vaccination again COVID-19 in the UK.^23-26^ Two studies assessed impacts of the relaxation of social restriction after vaccination,^24 25^ and one study assessed impacts of vaccination on hospital admissions,^26^ but covered a shorter time horizon and did not consider waning of immunity. Another study of SARS-CoV-2 vaccination in the UK focused on economic evaluations.^26^ Compared with previous studies, our study has separately considered the durability of naturally acquired and vaccine induced immunity, over a 10 year period from 2020 to 2029, and compared a wider range of plausible scenarios. We explicitly investigated the impacts of different types of immune responses to SARS-CoV-2 infection and vaccines on the COVID-19 epidemic in a country. Findings from our study will improve the understanding of key immunological parameters relevant to future changes in SARS-CoV-2 transmission dynamics and vaccination strategies.

Evidence from randomised controlled trials showed that vaccines against SARS-CoV-2 are efficacious in reducing symptomatic COVID-19 cases, compared with placebo.^17^ The reduction in symptomatic cases in the vaccine group may be attributable to infection protection or disease reduction. Available evidence showed that vaccines reduced the risk of SARS-CoV-2 virus infection in vaccinated individuals,^27^ and household members of vaccinated healthcare workers has a lower risk of COVID-19 infection than those of unvaccinated.^28^ In this study, we explored the impacts of different proportions of a vaccine’s infection protection efficacy and disease reduction efficacy. Different combinations of the two efficacy components have impacts on the transmission dynamics, depending on the duration of immune response and reinfectivity after waning of sterilising immunity. Because of lack of data, we assumed that vaccine efficacy is equally attributable to the infection and disease reduction immunity.

We assumed a range of overall vaccine efficacy, including 50%, 70% and 90%, without considering doses of vaccines. As most vaccinated individuals will eventually receive the second dose of vaccine, we might have under-estimated vaccine efficacy. On the other hand, vaccines that are efficacious against current SARS-CoV-2 virus may become less efficacious against new virus variants. Although the impact of new virus variants on the vaccine efficacy was not explicitly considered, the analysis included scenarios with a low vaccine efficacy (50%). In addition, we assume that there will be no important safety issues for vaccines licensed to use. We focused on the impacts and interactions of vaccine efficacy, different types of immune response to SARS-CoV-2, and assumed no more restrictions by NPI measures after return to normality in England from June 2021. Therefore, the pessimistic scenarios in our analyses may not be allowed to happen in the real world, as NPI (including lockdown) measures may be introduced again if the vaccination programmes are insufficient to avoid the new outbreaks of COVID-19.

This is a deterministic simulation model, and uncertainty in estimated parameters may have not been fully accommodated. For simplicity, stochastic uncertainty, to quantify confidence intervals around the model’s outputs, was not modelled. However, the model has been verified based on historically observed outcome data in England,^7^ and a large number of projection scenarios are explored. Although many uncertainties remain, including durability and types of naturally acquired and vaccine-induced immunity, our model can be updated to assess vaccination strategies, as new evidence emerges.

## Conclusions

Under optimistic scenarios, mass immunisation using efficacious vaccines may enable society safely to return to normality. However, under plausible scenarios with low vaccine efficacy and short durability of immunity, COVID-19 could continue to cause recurrent waves of severe morbidity and mortality despite frequent vaccinations, and necessitate stringent NPI restrictions. It is crucial to monitor the vaccination effects in the real world, and to better understand characteristics of naturally acquired and vaccine induced immunity against SARS-CoV-2.

## Data Availability

All data relevant to the study are included in the article or uploaded as online supplementary information.

## Contributors

FS designed, developed the model, retrieved data for estimating parameters, conducted computational calculations, and prepared the draft manuscript. MOB provided methodological support, helped interpret results and critically revised the draft manuscript. FS accepts full responsibility for the work and/or the conduct of the study, had access to the data, and controlled the decision to publish.

## Patient consent for publication

Not required.

## Ethics approval statement

Not applicable. No human participants involved in this modelling study using data from published or openly accessible sources.

## ONLINE SUPPLEMENTARY MATERIALS

**Supplementary Files:** Model structure, data sources, model parameters, and mathematical equations

**Supplementary Tables:** Additional results of relevant scenarios

## SUPPLEMENTARY TABLES

**Supplementary Table 1:** Additional outcomes for scenarios in Figure 3. Estimated numbers of deaths from COVID-19

| **Supplementary Table 1:** |  |  |  |  |  |  |  |  |  |
| --- | --- | --- | --- | --- | --- | --- | --- | --- | --- |
| Scenario | Vaccination:<br>waves (interval) | Overall<br>vaccine<br>efficacy | Reduction in<br>reinfectivity | Immunity<br>duration (days) |  | COVID-19 deaths |  |  |  |
|  |  |  |  | Natural | Vaccine | 2020-2021 | 2022-2025 | 2026-2029 | Total:<br>2020-2029 |
| Fig.3A | 1 | 0.9 | 0.6 | 730 | 365 | 112,878 | 140,312 | 4,871 | 258,061 |
| Fig.3B | 3(365) | 0.9 | 0.6 | 730 | 365 | 112,878 | - | 60,448 | 173,326 |
| Fig.3C | 5(365) | 0.9 | 0.6 | 730 | 365 | 112,878 | - | 752 | 113,630 |
| Fig.3D | 1 | 0.9 | 0.6 | 730 | 182 | 112,878 | 95,441 | 98,177 | 306,496 |
| Fig.3E | 3(365) | 0.9 | 0.6 | 730 | 182 | 112,878 | 12,372 | 52,244 | 177,494 |
| Fig.3F | 5(365) | 0.9 | 0.6 | 730 | 182 | 112,878 | - | 51,946 | 164,824 |

**Supplementary Table 2:** The projected numbers of COVID-19 deaths under various simulation scenarios

| Scenario | Vaccination:<br>waves (interval) | Overall<br>vaccine<br>efficacy | Reduction in<br>reinfectivity | Immunity<br>duration (days) |  | COVID-19 deaths |  |  |  |
| --- | --- | --- | --- | --- | --- | --- | --- | --- | --- |
|  |  |  |  | Natural | Vaccine | 2020-2021 | 2022-2025 | 2026-2029 | Total:<br>2020-2029 |
| st1-01 | 2(365) | 90% | 60% | 730 | 730 | 112,815 | - | - | 112,815 |
| st1-02 | 2(365) | 90% | 60% | 730 | 365 | 112,815 | - | - | 112,815 |
| st1-03 | 2(365) | 90% | 60% | 730 | 182 | 112,815 | - | - | 112,815 |
| st1-04 | 3(365) | 90% | 60% | 365 | 365 | 112,870 | - | - | 112,870 |
| st1-05 | 3(365) | 90% | 60% | 365 | 182 | 112,870 | - | - | 112,870 |
| st1-06 | 5(365) | 90% | 40% | 730 | 730 | 112,878 | - | - | 112,878 |
| st1-07 | 6(365) | 90% | 40% | 730 | 365 | 112,878 | - | - | 112,878 |
| st1-08 | 9(365) | 90% | 40% | 730 | 182 | 112,878 | - | - | 112,878 |
| st1-09 | 7(365) | 90% | 40% | 365 | 365 | 112,986 | - | - | 112,986 |
| st1-10 | 9(365) | 90% | 40% | 365 | 182 | 112,986 | - | - | 112,986 |
| st1-11 | 7(365) | 90% | 20% | 730 | 730 | 112,942 | - | - | 112,942 |
| st1-12 | 9(365) | 90% | 20% | 730 | 365 | 112,942 | - | 1,409 | 114,351 |
| st1-13 | 9(365) | 90% | 20% | 730 | 182 | 112,942 | 24,542 | - | 137,484 |
| st1-14 | 9(365) | 90% | 20% | 365 | 365 | 113,147 | 17,147 | 3,419 | 133,713 |
| st1-15 | 9(365) | 90% | 20% | 365 | 182 | 113,616 | 75,130 | 48,955 | 237,701 |
| st1-16 | 2(365) | 70% | 60% | 730 | 730 | 114,065 | - | - | 114,065 |
| st1-17 | 3(365) | 70% | 60% | 730 | 365 | 114,065 | - | - | 114,065 |
| st1-18 | 3(365) | 70% | 60% | 730 | 182 | 114,065 | - | - | 114,065 |
| st1-19 | 3(365) | 70% | 60% | 365 | 365 | 114,223 | - | - | 114,223 |
| st1-20 | 3(365) | 70% | 60% | 365 | 182 | 114,228 | 1,554 | - | 115,782 |
| st1-21 | 6(365) | 70% | 40% | 730 | 730 | 114,271 | - | - | 114,271 |
| st1-22 | 8(365) | 70% | 40% | 730 | 365 | 114,271 | - | - | 114,271 |
| st1-23 | 9(365) | 70% | 40% | 730 | 182 | 114,275 | 3,740 | 11,182 | 129,197 |
| st1-24 | 9(365) | 70% | 40% | 365 | 365 | 114,814 | 24,220 | 30 | 139,064 |
| st1-25 | 9(365) | 70% | 40% | 365 | 182 | 115,419 | 60,365 | 32,732 | 208,516 |
| st1-26 | 9(365) | 70% | 20% | 730 | 730 | 114,594 | 4,879 | 8,845 | 128,318 |
| st1-27 | 9(365) | 70% | 20% | 730 | 365 | 114,594 | 24,982 | 24,386 | 163,962 |
| st1-28 | 9(365) | 70% | 20% | 730 | 182 | 115,076 | 55,193 | 29,983 | 200,252 |
| st1-29 | 9(365) | 70% | 20% | 365 | 365 | 116,681 | 133,875 | 65,610 | 316,166 |
| st1-30 | 9(365) | 70% | 20% | 365 | 182 | 119,965 | 280,585 | 123,846 | 524,396 |
| st1-31 | 3(365) | 50% | 60% | 730 | 730 | 115,414 | - | - | 115,414 |
| st1-32 | 3(365) | 50% | 60% | 730 | 365 | 115,414 | - | - | 115,414 |
| st1-33 | 3(365) | 50% | 60% | 730 | 182 | 115,419 | - | - | 115,419 |
| st1-34 | 9(365) | 50% | 60% | 365 | 365 | 115,723 | 1,440 | - | 117,163 |
| st1-35 | 9(365) | 50% | 60% | 365 | 182 | 115,836 | 9,564 | - | 125,400 |
| st1-36 | 9(365) | 50% | 40% | 730 | 730 | 115,965 | 435 | 293 | 116,693 |
| st1-37 | 9(365) | 50% | 40% | 730 | 365 | 115,965 | 1,500 | 17,798 | 135,263 |
| st1-38 | 9(365) | 50% | 40% | 730 | 182 | 116,276 | 31,849 | 17,791 | 165,916 |
| st1-39 | 9(365) | 50% | 40% | 365 | 365 | 117,753 | 83,968 | 34,605 | 236,326 |
| st1-40 | 9(365) | 50% | 40% | 365 | 182 | 118,845 | 118,689 | 79,225 | 316,759 |
| st1-41 | 9(365) | 50% | 20% | 730 | 730 | 117,545 | 56,380 | 39,664 | 213,589 |
| st1-42 | 9(365) | 50% | 20% | 730 | 365 | 117,545 | 66,524 | 55,405 | 239,474 |
| st1-43 | 9(365) | 50% | 20% | 730 | 182 | 118,740 | 153,640 | 3,575 | 275,955 |
| st1-44 | 9(365) | 50% | 20% | 365 | 365 | 126,982 | 267,930 | 183,798 | 578,710 |
| st1-45 | 9(365) | 50% | 20% | 365 | 182 | 137,272 | 396,172 | 220,575 | 754,019 |

Notes to supplementary Table 2.
1. Assumptions: vaccine efficacy equally attributable to infection and disease protection; mass vaccination with 80% coverage of individuals aged 16+.
2. Vaccination waves (interval): number of waves and days between the starting dates of the two adjacent waves. This appendix table shows results of repeated annual mass vaccination programmes. There are maximally nine waves of annual mass vaccination programmes during 2021-2029. Under some scenarios, COVID-19 outbreaks before the end of 2029 can be prevented with 2-7 waves of annual vaccination programmes.

**Supplementary Table 3:** Results of sensitivity analyses. Estimated numbers of deaths from COVID-19

| Supplementary Table 3: |  |  |  |  |  |  |  |  |  |
| --- | --- | --- | --- | --- | --- | --- | --- | --- | --- |
| Scenario | Vaccination:<br>waves (interval) | Overall<br>vaccine<br>efficacy | Reduction in<br>reinfectivity | Immunity<br>duration (days) |  | COVID-19 deaths |  |  |  |
|  |  |  |  | Natural | Vaccine | 2020-2021 | 2022-2025 | 2026-2029 | Total:<br>2020-2029 |
| Different frequency and days between vaccination programmes |  |  |  |  |  |  |  |  |  |
| L92c1 | 2(730) | 0.9 | 60% | 730 | 730 | 112,815 | - | - | 112,815 |
| Lm92c1 | 2(730) | 0.9 | 60% | 730 | 365 | 112,815 | - | - | 112,815 |
| L94c2 | 4(730) | 0.9 | 40% | 730 | 730 | 112,878 | - | - | 112,878 |
| Ls916s3 | 16(182) | 0.9 | 40% | 730 | 182 | 112,942 | - | - | 112,942 |
| Ms93s1 | 3(182) | 0.9 | 60% | 365 | 182 | 112,870 | - | - | 112,870 |
| Ms913s2 | 13(182) | 0.9 | 40% | 365 | 182 | 112,878 | - | - | 112,878 |
| M916s3 | 16(182) | 0.9 | 20% | 365 | 365 | 113,145 | - | - | 113,145 |
| Ms918s3 | 18(182) | 0.9 | 20% | 365 | 182 | 113,153 | 5,724 | 4,569 | 123,446 |
| L73c1 | 3(730) | 0.7 | 60% | 730 | 730 | 114,065 | - | - | 114,065 |
| Lm73c1 | 3(730) | 0.7 | 60% | 730 | 365 | 114,065 | - | - | 114,065 |
| Ls710s2 | 10(182) | 0.7 | 40% | 730 | 182 | 114,275 | - | - | 114,275 |
| L718s3 | 18(182) | 0.7 | 20% | 730 | 730 | 114,576 | - | - | 114,576 |
| Ls718s3 | 18(182) | 0.7 | 20% | 730 | 182 | 114,600 | - | 13,802 | 128,402 |
| Lm718s3 | 18(182) | 0.7 | 20% | 730 | 365 | 114,576 | - | - | 114,576 |
| Ms73s1 | 3(182) | 0.7 | 60% | 365 | 182 | 114,228 | - | - | 114,228 |
| M718s2 | 18(182) | 0.7 | 40% | 365 | 365 | 114,642 | - | - | 114,642 |
| Ms718s2 | 18(182) | 0.7 | 40% | 365 | 182 | 114,674 | 650 | 5,067 | 120,391 |
| M718s3 | 18(182) | 0.7 | 20% | 365 | 365 | 115,808 | 24,794 | 19,794 | 160,396 |
| Ms718s3 | 18(182) | 0.7 | 20% | 365 | 182 | 116,168 | 64,685 | 47,241 | 228,094 |
| No reduction in reinfectivity after immunity waning |  |  |  |  |  |  |  |  |  |
| L99m0 | 9(365) | 0.9 | 0% | 730 | 730 | 113,012 | - | 1,255 | 114,267 |
| Lm99m0 | 9(365) | 0.9 | 0% | 730 | 365 | 113,012 | 17,702 | 7,512 | 138,226 |
| Ls99m0 | 9(365) | 0.9 | 0% | 730 | 182 | 113,070 | 30,619 | 47,229 | 190,918 |
| M99m0 | 9(365) | 0.9 | 0% | 365 | 365 | 113,684 | 52,806 | 35,665 | 202,155 |
| Ms99m0 | 9(365) | 0.9 | 0% | 365 | 182 | 115,880 | 261,365 | 29,700 | 406,945 |
| L79m0 | 9(365) | 0.7 | 0% | 730 | 730 | 115,522 | 49,095 | 28,213 | 192,830 |
| Lm79m0 | 9(365) | 0.7 | 0% | 730 | 365 | 115,522 | 51,428 | 56,044 | 222,994 |
| Ls79m0 | 9(365) | 0.7 | 0% | 730 | 182 | 117,170 | 167,268 | 499 | 284,937 |
| M79m0 | 9(365) | 0.7 | 0% | 365 | 365 | 123,398 | 250,649 | 159,941 | 533,988 |
| Ms79m0 | 9(365) | 0.7 | 0% | 365 | 182 | 143,421 | 381,311 | 350,640 | 875,372 |

## SUPPLEMENTARY FILES

### 1. Model structure and status

This is a discrete-time population dynamic simulation model. Population in England are partitioned into discrete categories by sex (male or female), age (0-4, 5-9, then by 10 year age bands, and 80+), and Covid-19 infection categories. The main infection categories include: susceptible (SU), exposed (EX), infected (IN), and recovered (RE) (appendix figure 1). The infected individuals are further categorised as asymptomatic, symptomatic, self-isolated, and hospitalised.

**Appendix figure 1:**
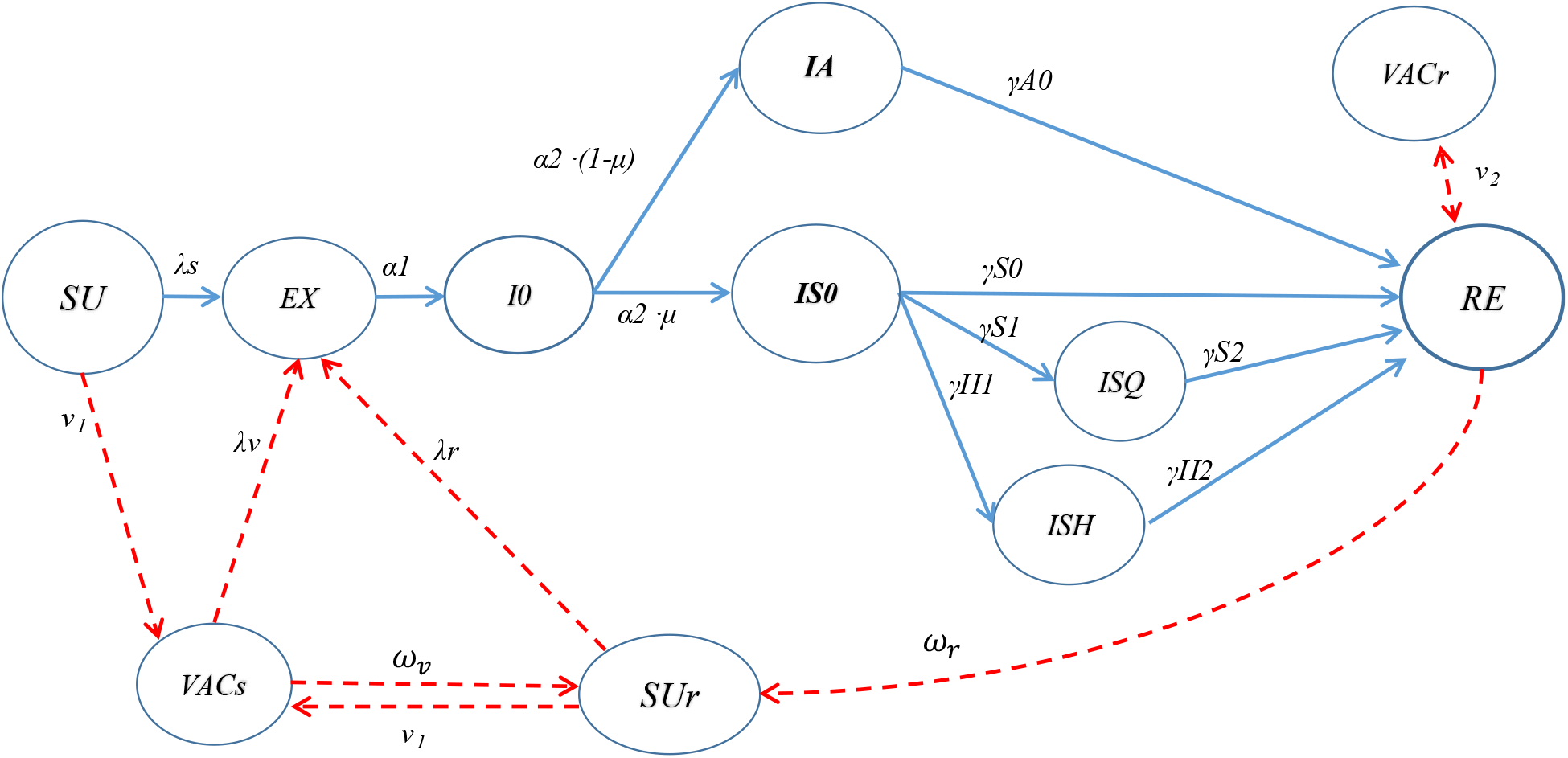
Model structure and transmission across status. **Definitions of compartmental variables in appendix figure 1** • SU: susceptible individuals • SUr: Individuals susceptible to reinfection after immunity waning • EX: exposed individuals, not infectious • I0: infectious, before symptom onset • IA: infectious individuals with no or very mild symptoms • IS0: symptomatic patients who are not quarantined • ISQ: symptomatic patients self-isolated • ISH: symptomatic patients who are hospitalised • RE: recovered from covid-19 infection • VACs: vaccinated susceptible individuals • VACr: vaccinated individuals who recovered from infection **Transition parameters in appendix figure 1:** • λ_s_: Force of infection (*λ*) measures the risk (probability) of infection transmission, which is a function of transmission rate (β) and the prevalence of infectious individuals (*I*) among the population (*N*): *λ=β·I/N*.^1^ • β: The transmission rate β in this discrete-time model is defined as the average number of individuals infected daily by an infectious person. It is a function of the number of daily close contacts per person (c), and the transmission risk per contact between a susceptible and an infectious individual (η): i.e., *β=c·η*. ^2^ • α1: rate of progressing from being exposed to being infectious. • α2: rate of progressing from being asymptomatic infectious to symptomatic. • *μ*: proportion of infected individuals who will be symptomatic; age-specific • *infA*: The fraction of infection force for infected individuals with no or mild symptoms. It was assumed that infA=0.5 in this study. • fS0: fraction of symptomatic patients who will not be quarantined. • fSq: fraction of symptomatic patients who will be quarantined (self-isolation). • fSh: fraction of symptomatic patients who will be hospitalised (including ICU admission). • γA0: rate of recovering for asymptomatic individuals • γS0 rate of recovering for symptomatic, not isolated/hospitalised patients • γS1: rate of being isolated in symptomatic patients • γS2: rate of recovering in isolated patients • γH1: rate of being hospitalised for symptomatic patients • γH2: rate of recovering in hospitalised patients • ν_1_: rate of vaccinating susceptible individuals • ν_2_: rate of vaccinating recovered individuals • *ⴍ*_v_: rate of immunity waning in vaccinated individuals • *ⴍ*_r_: rate of immunity waning in recovered individuals

All individuals in England are assumed to be susceptible to SARS-CoV-2 infection at the beginning of 2020. Susceptible individuals may be infected by contacting infectious individuals, and the infection status is changed from “susceptible” (SU) to “exposed” (EX). The exposed individuals are not infectious during the early incubation period, but start to be infectious before the onset of symptoms. Individuals infected with SARS-CoV-2 virus may have no or very mild symptoms (IA), and palpable symptoms (symptomatic or clinical infections). Asymptomatic individuals can spread SARS-CoV-2 virus before recovery, although the transmission risk may be lower than symptomatic patients. Symptomatic patients are further classified into three categories: symptomatic patients who are neither isolated nor hospitalised (IS0), those who self-isolate at home (ISQ), and those who are hospitalised (including those being admitted to intensive care units) (ISH). Symptomatic patients are infectious and can transmit the virus to susceptible people before being isolated, hospitalised or recovered. We assume that hospitalised patients (ISH) are well isolated and no longer able to spread the virus to the susceptible population, although infectious patients who are self-isolated at home (ISQ) may transmit virus to household contacts.

Individuals may recover from previous infection of SARS-CoV-2 (RE), and the susceptible and recovered individuals may be vaccinated with vaccines again SARS-CoV-2 virus (VAC1 and VAC2). Individuals recovered or effectively vaccinated may develop immune responses against infection with SARS-CoV-2. However, if the protective immunity is not long lasting, individuals who have recovered or vaccinated may become susceptible again after the waning of the immunity (SUr).

The immune response against COVID-19, either by naturally acquired from past SARSE-CoV-2 infection or vaccine-induced, may be long lasting or short-lived. Immune response may reduce susceptibility of individuals to infection (infection protection, or sterilising, immunity), reduce disease severity after being infected (disease reduction immunity), and reduce infectivity of those who are reinfected after recovery or being vaccinated (reinfectivity reduction immunity).^3^ According to existing evidence on immunological characteristics for other human coronaviruses, immunity against reinfection (sterilising immunity) may be waning in several months, while disease and reinfectivity reduction responses are likely long lasting.^3^ According to these basic concepts specified by Lavine et al,^3^ we incorporate the three types of immune responses into the model, to explicitly evaluate their impacts on future transmission dynamics (appendix figure 2).

**Appendix figure 2:**
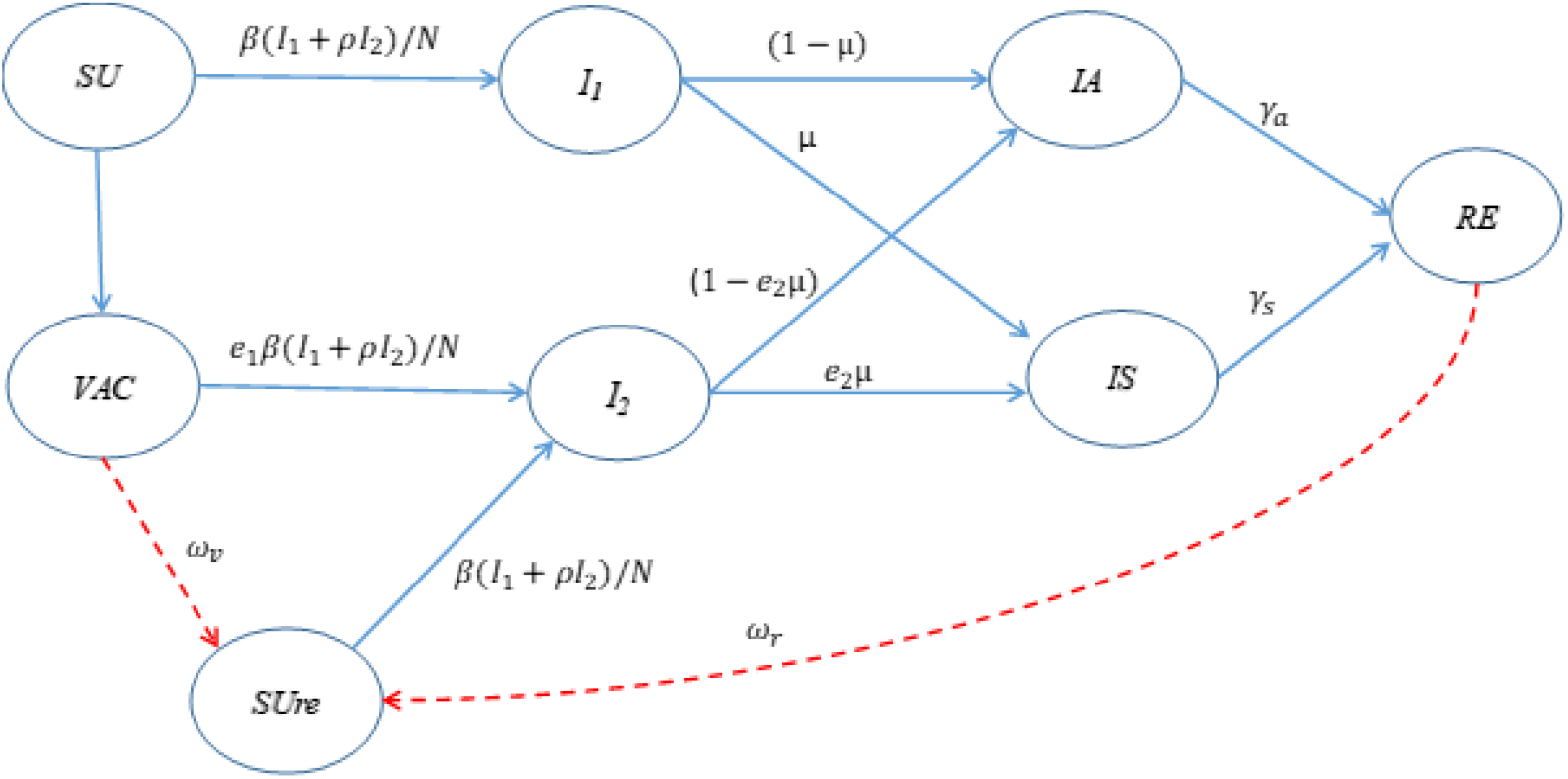
Types of immune responses by natural infection or vaccination. **Notes to appendix figure 2:** • Appendix figure2 is a simplified version of appendix figure 1, not showing isolation and hospitalisation for symptomatic patients. • *N*: The number of the population • β: The transmission rate, i.e., the average number of individuals infected daily by an infectious person. It is a function of the number of daily close contacts per person (c), and the transmission risk per contact between a susceptible and an infectious individual (η): i.e., *β=c·η*. • *I*_*1*_: Infectious individuals with primary infection • *I*_*2*_: Infectious individuals with secondary infection (infected after being vaccinated or recovered) • ρ: Relative infectivity of the secondary infection (*I*_*2*_) compared with the primary infection (*I*_*1*_). For example, if ρ=0.6, the infectivity of *I*_2_ is 40% lower than the infectivity of *I*_*1*_ • *μ*: proportion of infected individuals who will be symptomatic; age-specific • *e*_1_: Relative efficacy of vaccine for sterilising immunity, reducing risk of virus transmission • *e*_*2*_: Relative efficacy of vaccine for pathology reduction, reducing the proportion of symptomatic cases after being infected • *IA*: Asymptomatic individuals • *IS*: Symptomatic patients • *γ*_*a*_ : Average rate of recovering of asymptomatic individuals • *γ*_*s*_ : Average rate of recovering of asymptomatic individuals • *ⴍ*_v_: rate of immunity waning in vaccinated individuals • *ⴍ*_r_: rate of immunity waning in recovered individuals

#### Overall and partial vaccine efficacy

Results of randomised controlled trials shown that vaccines may be >90% efficacious (e.g., Pfizer mRNA vaccine) in reducing severe symptomatic diseases, compared with the placebo group.^4^ In appendix figure 2, *e*_1_ and *e*_*2*_ are parameters of vaccine’s efficacy in blocking virus transmission and reducing symptomatic cases in the infected, respectively. The reduction in symptomatic cases in the vaccine group may be due to the prevention of infection in susceptible individuals (related to *e*_1_), or a lower proportion of infected individuals being symptomatic in the vaccine group (related to *e*_*2*_), or due to a combination of both. Let λ is the transmission risk and μ is the proportion of symptomatic cases in the infected without vaccination. After being vaccinated, the transmission risk is reduced to *λ·e_1_*, and the proportion of symptomatic cases reduced to *μ·e_2_*. For a vaccine with 90% efficacy in reducing the number of symptomatic cases (compared with the control group), it should be true that *e*_1_ *· e_2_ =(1-0*.*90)* or *e*_*1*_=*0*.*10/ e_2_*. There are many different possible combinations of *e*_1_ and *e*_*2*_ for a given overall efficacy in reducing symptomatic cases. For example, *e*_1_=*e*_*2*_=*SQRT(0*.*30)=0*.*548* corresponds to a 70% efficacy of vaccine with equal sterilising immunity and pathology reduction. If *e*_1_=1 (i.e., zero efficacy in sterilising immunity), all vaccine efficacy will be attributable to the pathology reduction, with *e*_*2*_=0.30, for a vaccine with 70% efficacy. The partial efficacy is calculated using: *Ei* = 1 − (1-*Eo*)/*Ed*, where *Ei* is the partial efficacy for infection protection, *Ed* is the partial efficacy for disease reduction, and *Eo* is the overall vaccine efficacy. The equal partial efficacy is calculated by: 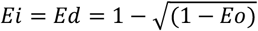. For vaccines with 90%, 70% and 50% overall efficacy, the equal partial efficacy for the infection protection and disease reduction is 69.4%, 45.2%, and 29.3%, respectively.

### 2. Parameterisation, data sources, and simulation scenarios

#### 2.1 TRANSITION PARAMETERS AND DISTRIBUTION OF INFECTIOUS PERIOD

In appendix figure 1, force of infection (*λ*) measures the risk of infection,^1^ which is a function of transmission rate (η) and the prevalence of existing infectious individuals (*I*) among the population (N): *λ*=*η*·*I*/*N*. The transmission rate *η* in the discrete-time model can be defined as the average number of new infected individuals generated daily by an infected person. That is, *η=Rt/T*, in which *Rt* is effective reproduction number and T is the average infectious period for infected individuals. We calculated *η* as a function of the number of daily contacts per person (c), and the risk of transmission per contact between a susceptible and an infected individual (β): η=c*β.^2^

The transition rate between model’s compartments in infectious models is often assumed to be constant, calculated by 1/x, in which “x” is the average period that subjects remain before the transition to the model’s next compartments.^1^ Therefore, the infectious period in standard SIR or SEIR models is usually assumed to be exponentially distributed, with some limitations of the use of exponentially distributed infectious period.^5 6^ In this study, we assumed that the transition probability between model’s compartments are based on gamma distributed period that individuals remain in a compartment.^7^ The transition probability (y) at t is: *y*_*t*_=(*cg*_*t*_ − *cg*_*t*−1_)/(1−*cg*_*t*_), where *cg*_*t*_ is the gamma cumulative probability by the end of t. Given mean and shape (k) parameters, the gamma distribution based transition probability is used as a deterministic value to estimate the number of individuals moving between two status in this study.

#### 2.2 PARAMETERISATION AND DATA SOURCES

We estimated initial parameters based on relevant literature and data from the UK government websites (appendix table 1). Key parameters were calibrated according to the reported numbers of covid-19 deaths, hospitalised patients, and the prevalence of infected individuals in England from January 2020 to January 2021.^8^

**Appendix table 1:**
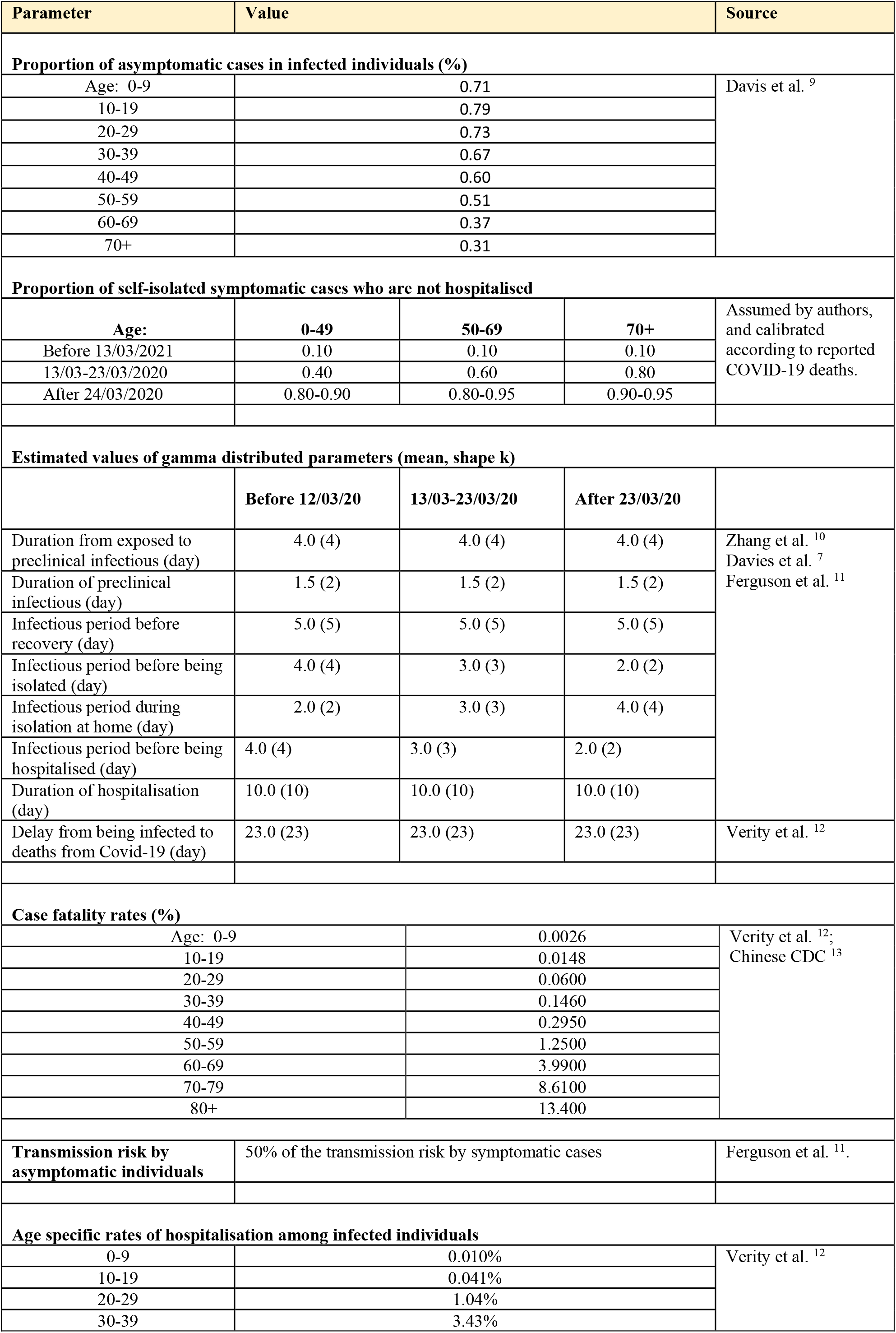
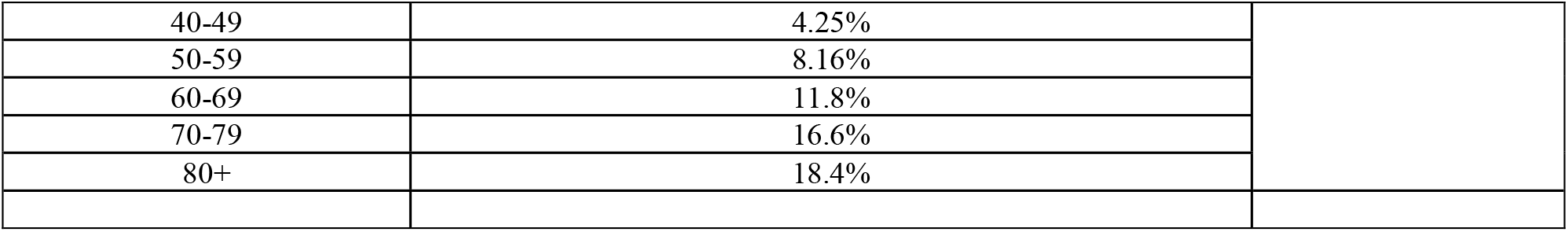
Summary of key model parameters.

| Parameter | Value |  |  | Source |
| --- | --- | --- | --- | --- |
| Proportion of asymptomatic cases in infected individuals (%) |  |  |  |  |
| Age: 0-9 | 0.71 |  |  | Davis et al. <sup>9</sup> |
| 10-19 | 0.79 |  |  |  |
| 20-29 | 0.73 |  |  |  |
| 30-39 | 0.67 |  |  |  |
| 40-49 | 0.60 |  |  |  |
| 50-59 | 0.51 |  |  |  |
| 60-69 | 0.37 |  |  |  |
| 70+ | 0.31 |  |  |  |
| Proportion of self-isolated symptomatic cases who are not hospitalised |  |  |  |  |
| Age: | 0-49 | 50-69 | 70+ | Assumed by authors, and calibrated according to reported COVID-19 deaths. |
| Before 13/03/2021 | 0.10 | 0.10 | 0.10 |  |
| 13/03-23/03/2020 | 0.40 | 0.60 | 0.80 |  |
| After 24/03/2020 | 0.80-0.90 | 0.80-0.95 | 0.90-0.95 |  |
| Estimated values of gamma distributed parameters (mean, shape k) |  |  |  |  |
|  | Before 12/03/20 | 13/03-23/03/20 | After 23/03/20 |  |
| Duration from exposed to preclinical infectious (day) | 4.0 (4) | 4.0 (4) | 4.0 (4) | Zhang et al. <sup>10</sup><br>Davies et al. <sup>7</sup><br>Ferguson et al. <sup>11</sup> |
| Duration of preclinical infectious (day) | 1.5 (2) | 1.5 (2) | 1.5 (2) |  |
| Infectious period before recovery (day) | 5.0 (5) | 5.0 (5) | 5.0 (5) |  |
| Infectious period before being isolated (day) | 4.0 (4) | 3.0 (3) | 2.0 (2) |  |
| Infectious period during isolation at home (day) | 2.0 (2) | 3.0 (3) | 4.0 (4) |  |
| Infectious period before being hospitalised (day) | 4.0 (4) | 3.0 (3) | 2.0 (2) |  |
| Duration of hospitalisation (day) | 10.0 (10) | 10.0 (10) | 10.0 (10) |  |
| Delay from being infected to deaths from Covid-19 (day) | 23.0 (23) | 23.0 (23) | 23.0 (23) | Verity et al. <sup>12</sup> |
| Case fatality rates (%) |  |  |  |  |
| Age: 0-9 | 0.0026 |  |  | Verity et al. <sup>12</sup>;<br>Chinese CDC <sup>13</sup> |
| 10-19 | 0.0148 |  |  |  |
| 20-29 | 0.0600 |  |  |  |
| 30-39 | 0.1460 |  |  |  |
| 40-49 | 0.2950 |  |  |  |
| 50-59 | 1.2500 |  |  |  |
| 60-69 | 3.9900 |  |  |  |
| 70-79 | 8.6100 |  |  |  |
| 80+ | 13.400 |  |  |  |
| Transmission risk by asymptomatic individuals | 50% of the transmission risk by symptomatic cases |  |  | Ferguson et al. <sup>11</sup>. |
| Age specific rates of hospitalisation among infected individuals |  |  |  |  |
| 0-9 | 0.010% |  |  | Verity et al. <sup>12</sup> |
| 10-19 | 0.041% |  |  |  |
| 20-29 | 1.04% |  |  |  |
| 30-39 | 3.43% |  |  |  |

|  |  |
| --- | --- |
| 40-49 | 4.25% |
| 50-59 | 8.16% |
| 60-69 | 11.8% |
| 70-79 | 16.6% |
| 80+ | 18.4% |

We obtained population statistics in England (estimates of mid-year 2020) from Office for National Statistics. It was assumed that all individuals in England were susceptible to SARS-CoV-2 infection at the beginning of 2020. By contacting with infectious individuals, susceptible individuals may be infected, and their infection category is changed from “susceptible” (SU) to “exposed” (EX). “Exposed” refers to the pre-infectious status of infected individuals. According to data from previous studies, the period of incubation before symptom onset was on average 5.5 days,^10^ and the exposed individuals start to be infectious about 1.5 days before the onset of symptoms.^7 11^ Therefore, we assumed a gamma distribution of incubation period, with a mean non-infectious period of 4 days (k=4.0) after being exposed, and a mean infectious period of 1.5 days (k=2) before symptom onset.

Individuals infected with SARS-CoV-2 virus may have no or very mild symptoms (asymptomatic infected), and palpable symptoms (symptomatic patients). As in previous modelling studies ^7 11^, it was assumed that asymptomatic individuals can spread SARS-CoV-2 virus before recovery, although the infectious risk was assumed to be half of symptomatic patients.^11^ We used age-specific rates of asymptomatic cases in the infected individuals, reported in a study based on data from 6 countries^9^ (appendix table 1).

Symptomatic patients are further classified into three categories: symptomatic patients who are neither isolated nor hospitalised (mainly at the initial phase of the epidemic), those who are self-isolated at home, those who are hospitalised (see appendix figure 1). We assume that asymptomatic individuals were not isolated, although the average number of daily contacts could be reduced by non-pharmaceutical interventions (NPIs), including social distancing, testing, contact tracing, and lockdown. Assumed proportions of self-isolation of symptomatic cases who are not hospitalised, depending on age and NPI measures are shown in appendix table 1.

We assume that only symptomatic patients are hospitalised, and age specific rates of hospitalisation among symptomatic individuals were from Verity et al.^12^ The hospitalisation rates were calibrated according to reported numbers of hospitalised patients with covid-19 in England.^14^ Based on the reported number of hospitalised patients and estimated number of symptomatic cases, the hospitalisation rate was estimated to be 70% lower than the estimated by Verity et al.^12^ Symptomatic patients are infectious and can transmit the virus to susceptible people before being hospitalised or isolated. We assume that hospitalised patients are no longer able to spread the virus to susceptible individuals in the community. However, infected individuals who are self-isolated at home may transmit virus to household contacts. The infectious period before recovery was assumed to be gamma distributed, with a mean value of 5 days. Before implementing any NPIs, the infectious period of symptomatic cases was of a mean value of 4 days (k=4) before being quarantined or hospitalised. After implementing NPI measures, the infectious period for isolated and hospitalised patients was reduced, having a mean value of 2 days (k=2). The mean hospital stay was assumed to be 10 days (k=10) (including ICU admitted patients) (appendix table 1). Verity et al estimated that the average duration from symptom onset to death was 17.8 days.^12^ Therefore, we assume that covid-19 related deaths occur on average 23 days (k=23) after being exposed/infected.

The simulation starts from 1 January 2020, over a period of 10 years until the end of 2029. We assume that the first exposed case was imported to England on January 15th 2020, and the daily number of infectious cases imported was increased by one until 9 February 2020, with a total number of 351 cases imported in 25 days. The sex-and-age-specific numbers of household and community daily contacts per person in the UK were obtained from a study in 8 European countries.^15 16^ For the purpose of simplicity, we considered only household contacts (relevant to self-isolation at home) and general daily contacts (for all types of contacts). The risk of positive transmission per contact between susceptible and infectious individuals (β) was estimated by calibrating estimated and reported numbers of covid-19 deaths in England, household and general daily contacts per person, and other model parameters. The transmission risk by asymptomatic individuals was assumed to be 50% by symptomatic patients.^11^

In this study, all COVID-19 related deaths are assumed to be from symptomatic cases, and age specific case fatality rates were based on a study by Verity et al.^12^ We assume that individuals infected with Covid-19 will not die from other causes before recovery. Average sex and age specific rates of all-cause deaths in England during 2015-2019 ^17^ were applied to people who are not infected with or recovered from covid-19. For simplicity and maintaining a stable population, we assumed that the number of births at day t equals to the number of all deaths at day t-1. Furthermore, for simplicity, we did not consider the influence of migration on the population. We adjusted the number of individuals belong to an age group (all <80+) at the beginning of the year since 2021 by shifting 20% (for age group 0-4 and 5-9) or 10% (for age group 10-19, … 70-79) of them to the adjacent higher age group.

##### NPI AND SEASONAL IMPACTS ON TRANSMISSION PARAMETERS

Since March 2020, NPI measures were recommended and gradually tightened in England, including hand washing, mouth covering when coughing in public places, home isolation of individuals with COVID-19 like symptoms, shielding of vulnerable individuals, avoiding non-essential contacts, and maintaining social distancing. These measures reduced contacts and transmission risk, and shortened the period of transmission by symptomatic individuals. We assumed that the general population’s contact rates were reduced by 10% to 40%, depending on age and co-morbidity (appendix table 2).

Based on the reported number of COVID-19 deaths, we estimate that the transmission risk per contact between infectious and susceptible individuals was reduced by 32%, from β=0.068 before the implementation of any NPIs to 0.046 by 15 March 2020. The UK government put lockdown measures in place from 24 March 2020, including working from home if possible, closure of schools and non-essential shops, pubs and restaurants, avoiding non-essential travelling, and cancelling gathering activities. We assume that numbers of general population contacts were reduced by 60-85%. We assume that the household contacts were not influenced by the NPI measures.

**Appendix table 2:** Assumed impacts of NPIs on general contacts in England. Notes: Values are scaling fractions to reduce the normal contacts. For example, a fraction of 0.80 means the contacts are reduced by 20%.

| Time | Age group |  |  |  |
| --- | --- | --- | --- | --- |
|  | 0-19 | 20-59 | 60-69 | 70+ |
| Before 13/3/2020 | 1.00 | 1.00 | 1.00 | 1.00 |
| 13/03/2020- | 0.90 | 0.80 | 0.70 | 0.60 |
| 17/03/2020- | 0.80 | 0.70 | 0.60 | 0.50 |
| 24/03/2020- | 0.40 | 0.30 | 0.20 | 0.15 |
| 05/07/2020- | 0.60 | 0.55 | 0.40 | 0.20 |
| 01/09/2020- | 0.80 | 0.60 | 0.50 | 0.30 |
| 05/11/2020- | 0.60 | 0.30 | 0.20 | 0.15 |
| 02/12/2020- | 0.70 | 0.60 | 0.50 | 0.30 |
| 05/01/2021- | 0.40 | 0.30 | 0.20 | 0.15 |
| 08/03/2021- | 0.80 | 0.60 | 0.50 | 0.30 |
| 01/06/2021- | 1.00 | 1.00 | 1.00 | 1.00 |

The lockdown measures in England started to be relaxed from 13 May 2020 by allowing partial returning to work. Further relaxing of control measures followed, including reopening of some shops and allowing outdoor meetings up to six people from 1 June, re-opening of more non-essential shops from 5 June, and further relaxing of restrictions (such as re-opening of pubs and restaurants) from 5 July 2020. However, social distancing measures was maintained and face covering was required where social distancing could not be implemented. From 1 September 2020, schools in England were re-opening. Consequently, the transmission risk per contact between susceptible and infectious individuals was increased to 0.052 since September. The impacts of these changes in NPIs were reflected in the assumed social contacts and transmission risk. Because of the new virus variant in the UK,^18^ the average transmission risk per contact was increased to β=0.052 by November 2020, and β=0.056 by the end of 2020.

To incorporate the impact of seasonality on future projections since 2021, we assumed that the transmission risk from April to September is 20% lower than that from October to February, according to observed changes in transmission dynamics during March 2020 to January 2021 in England.

##### MODEL VALIDATION

We used the developed model and initially estimated parameters to simulate the covid-19 epidemic in England from January 2020 to January 2021. Key parameters were calibrated based on reported covid-19 related deaths, hospitalised patients, and infection rates in England.

We assume that the first exposed case was imported to England on 15 January 2020, and the number of cases imported each day increased by one more case than the previous day until 9 February 2020 (the total number of cases imported in 25 days was therefore 351). We don’t use the reproduction number (R0 or Rt) as an input parameter, but derived the reproduction numbers based on a method used by Giordano and colleagues (see equation 55 in Mathematical equations) ^1 19^. We estimated that the basic reproduction number (R0) was 3.68 at the initial stage of the COVID-19 epidemic before any control measures were taken in England, which is similar to findings from previous studies.^7 11^ Following the implementation of NPI measures, the estimated reproduction value (Rt) was reduced to 0.66 by 24 March. The Rt value was increased to 0.92 by 5 July 2020 after the NPI measures were relaxed, and Rt was about 1.13 after school reopening since September 2020. The Rt was reduced to 0.75 since 5 November 2020 after reintroducing NPI measures, increased to about 1.19 after relaxing NPIs since 2 December 2020, and reduced again to about 0.60 since 5 January 2021 after reintroducing lockdown measures (plus rolling out of vaccination) (appendix figure 3). The estimated R values were within the range of the reported in England (https://coronavirus.data.gov.uk/).

**Appendix figure 3.**
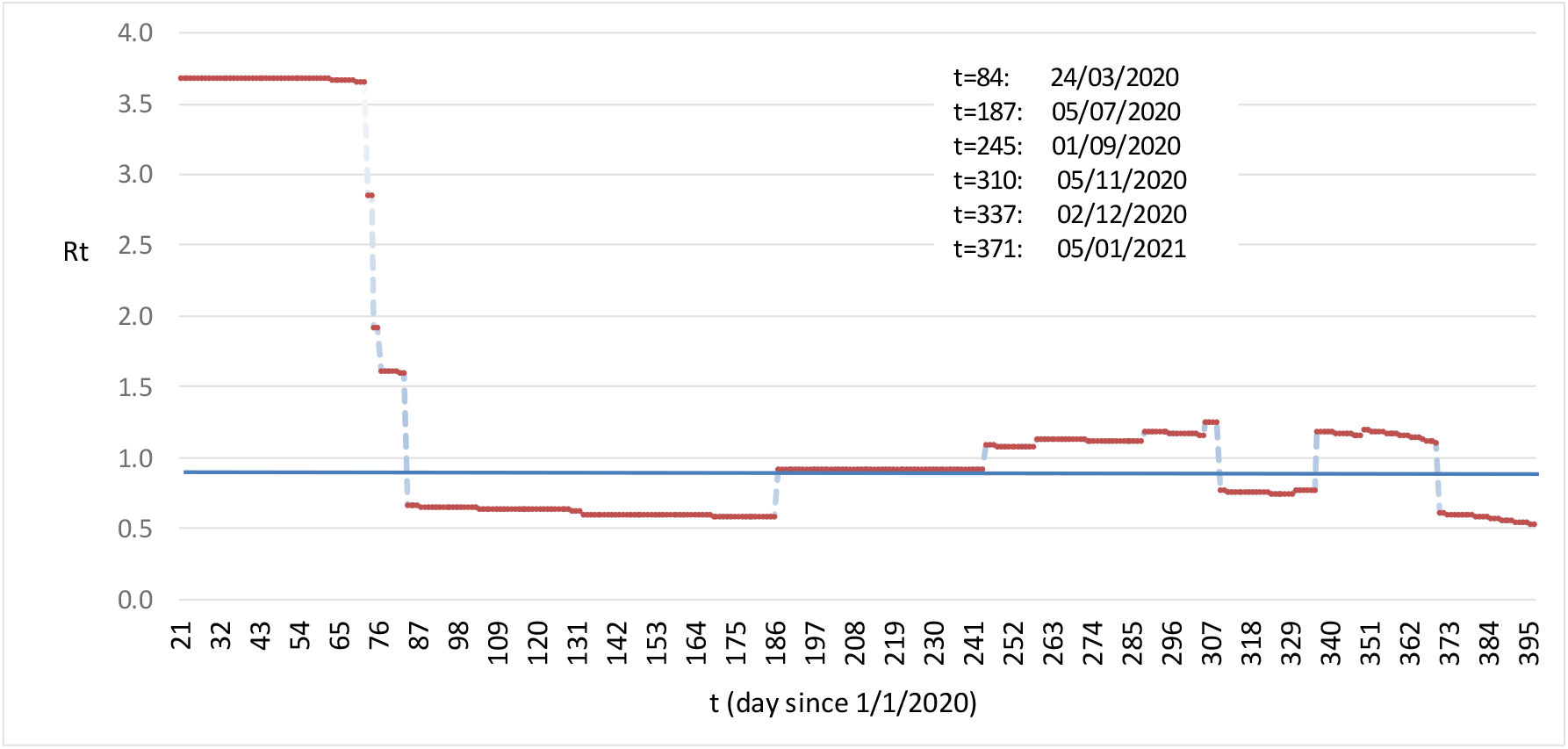
Estimated reproduction numbers, between 01/2020 – 01/2021, in England.

The model estimated that the prevalence of the recovered was 5.5 by 26 April, 7.6% by 24 May, and 8.5% by 24 June 2020, which were similar to the estimated rates of positive antibodies to Covid-19 in the UK (i.e., 7.1% in May-June 2020).^20^ Data on the prevalence of infected individuals in the community was available from May 2020. The model estimated prevalence of infected individuals from January 2020 to January 2021, which had a similar trend as the reported prevalence in England (appendix figure 4).

**Appendix figure 4:**
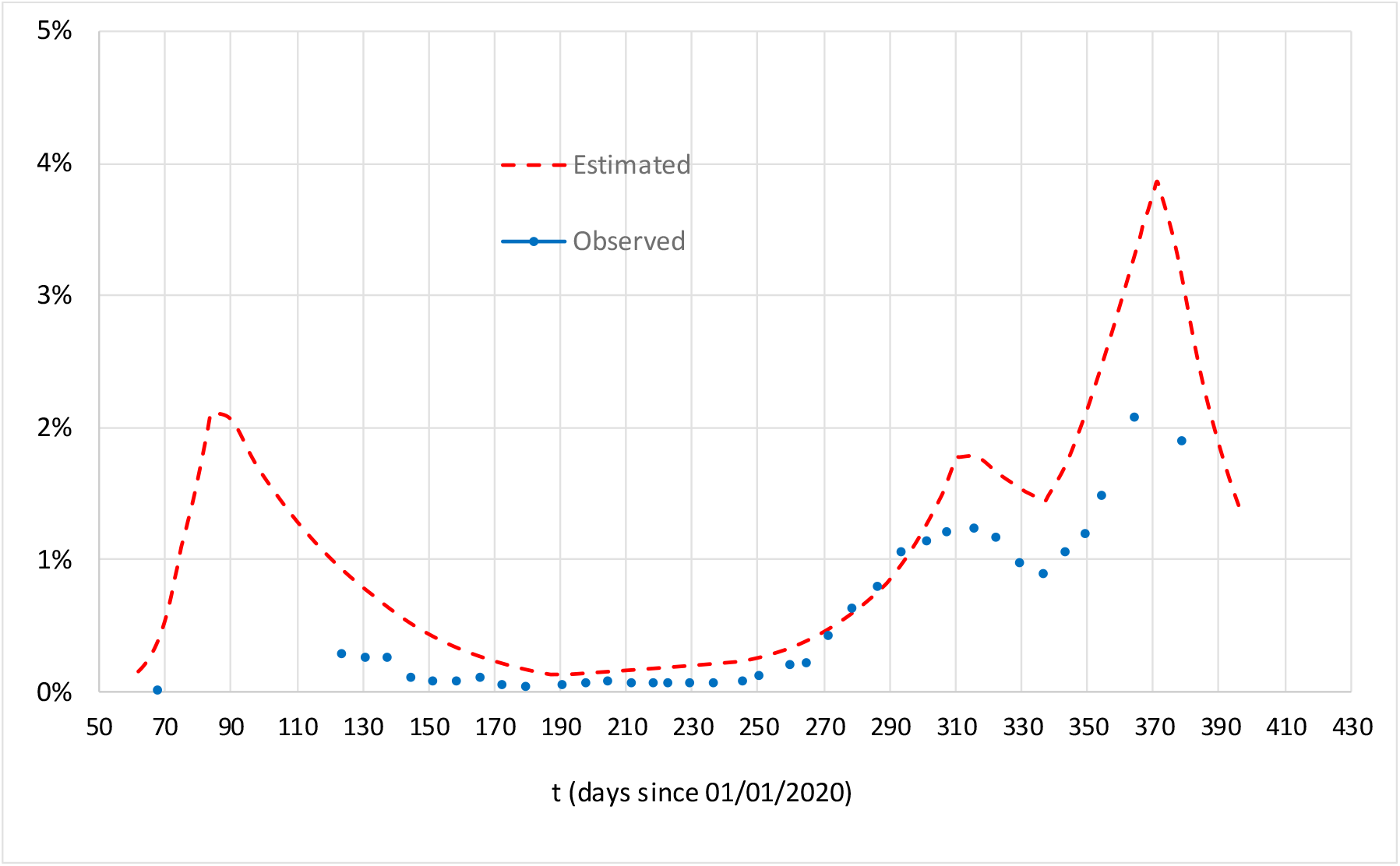
Estimated and reported prevalence of infection, from January 2020 to January 2021, in England.

Changes in the estimated numbers of hospitalised COVID-19 patients were of similar trends as the reported numbers of hospitalised patients during 01/2020-01/2021. However, there were considerable differences at peak time points (appendix figure 5), which may be explained by reduced hospitalisation rates during peak period.

**Appendix figure 5:**
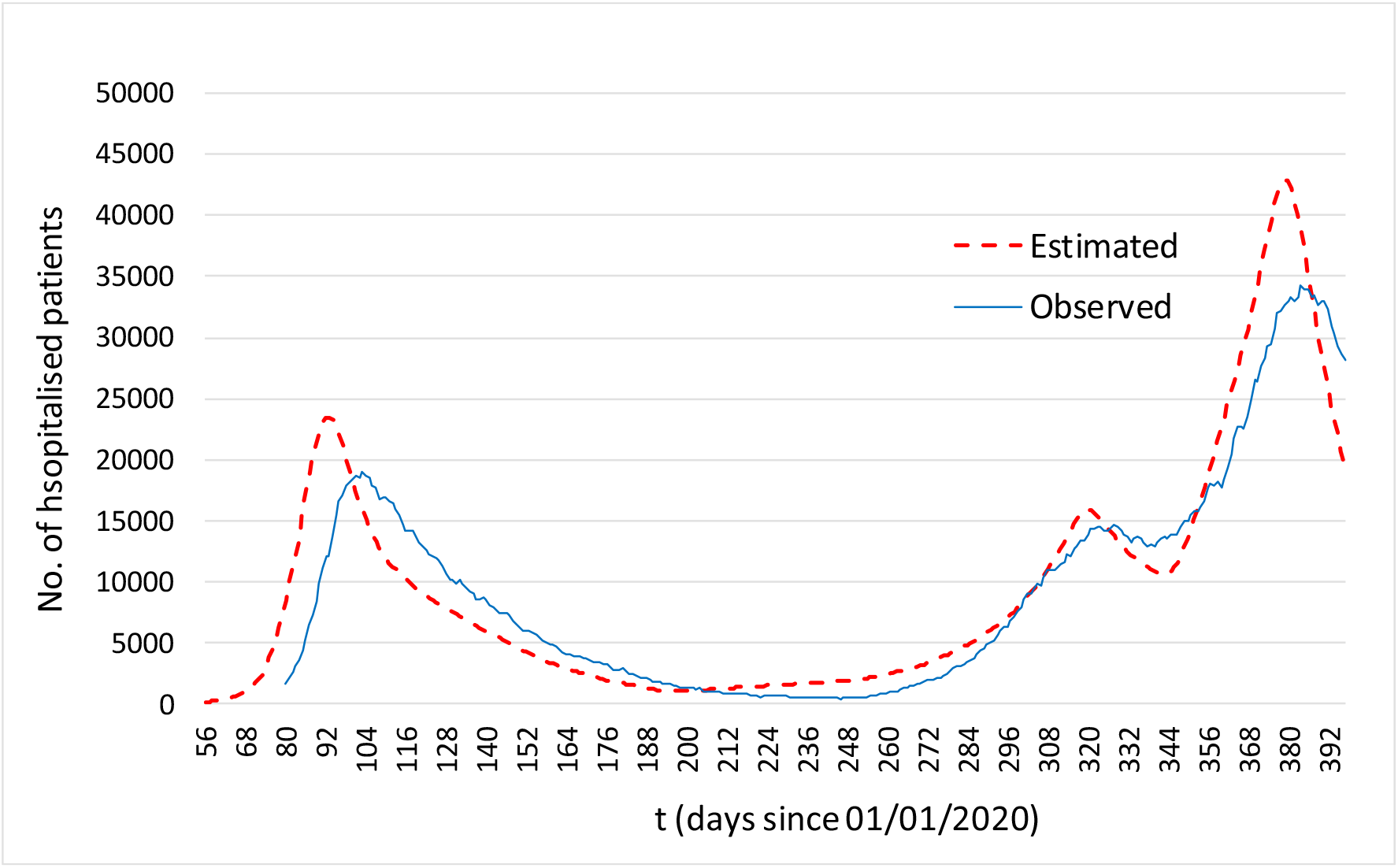
Reported and estimated numbers of hospitalised Covid-19 patients, during 01/2020-01/2021, in England.

Appendix figure 6 shows that the estimated daily deaths well matched the observed daily deaths from Covid-19, from January 2020 to January 2021, in England.

**Appendix figure 6:**
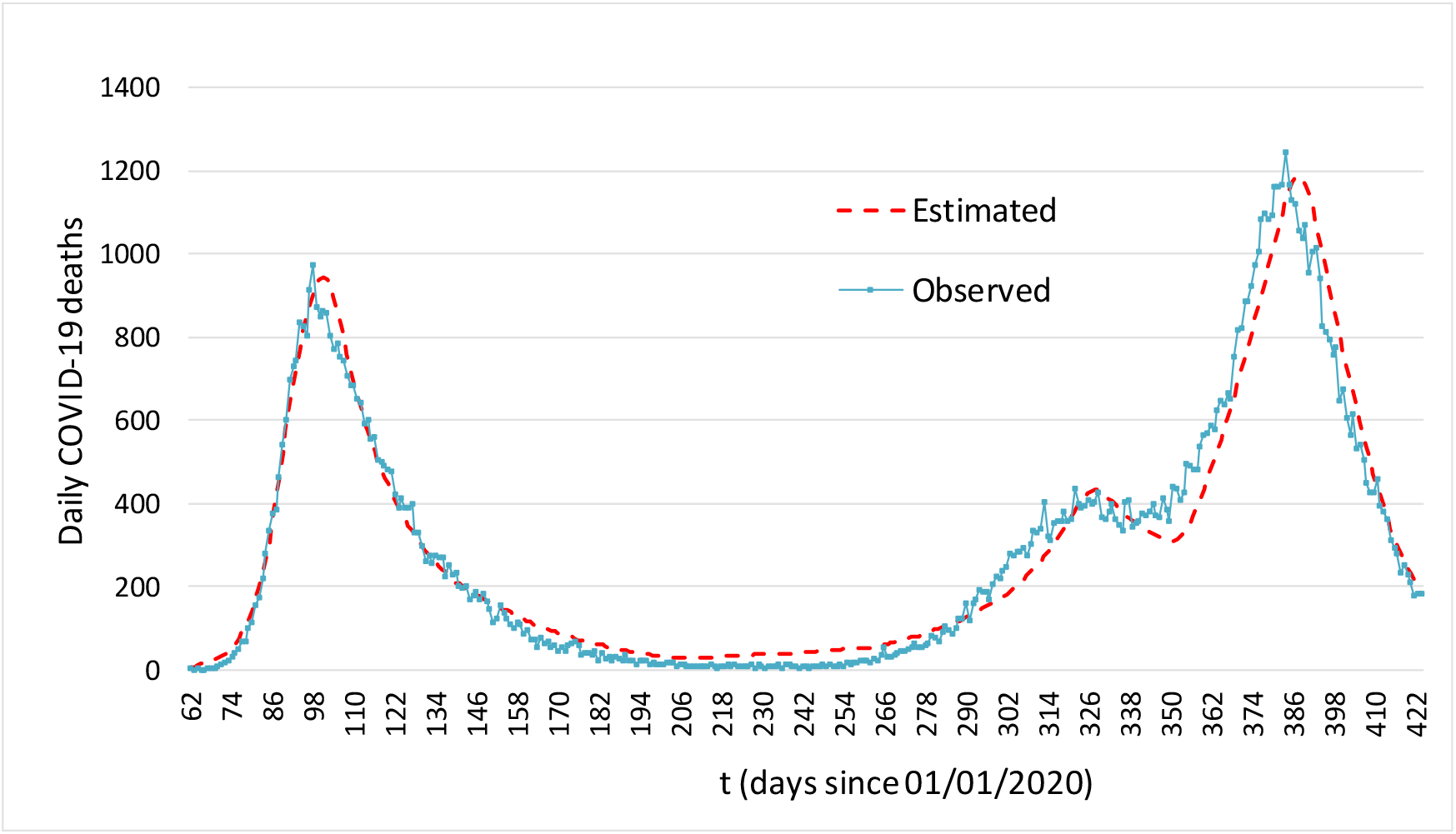
Reported and estimated daily deaths from Covid-19, during 01/2020-01/2021, in England.

### 3. Vaccination and projection scenarios

We used estimates of transmission parameters, age-specific hospitalisation rates and case fatality rates in January 2021 to project COVID-19 deaths from 2021-2029, under various scenarios of vaccine efficacy, durability of both naturally acquired and vaccine induced immunity, and reduction in reinfectivity. We assume that, since June 2021, there are no more restrictions on social activities, and social contracts are return to normal as before the pandemic, although basic hygienic measures would be maintained. Projection scenarios are defined from the following aspects: vaccine efficacy, durability of natural and vaccine induced sterilising immunity, and reduction in reinfectivity after the waning of infection protection immunity. In this study, we assume that the disease reduction and reinfectivity reduction immunity are long lasting, according to immunological characteristics of other endemic HCoVs.^3^

Results of randomised controlled trials shown that vaccines may be >90% efficacious (e.g., Pfizer mRNA vaccine) in reducing severe symptomatic diseases, compared with the placebo group. Assume that *e*_1_ and *e*_*2*_ are parameters of vaccine’s efficacy in blocking virus transmission and reducing symptomatic cases in the infected, respectively. The reduction in symptomatic cases in the vaccine group may be due to the prevention of infection in susceptible individuals (i.e., infection protection, related to *e*_1_), or a lower proportion of infected individuals being symptomatic in the vaccine group (i.e., disease reduction, related to *e*_*2*_), or due to a combination of both. For a vaccine with 90% efficacy in reducing the number of symptomatic cases (compared with the control group), it is true that *e*_1_*·e_2_=(1-0*.*90)*, or *e*_1_*=(1-0*.*90)/ e_2_*. There are many different possible combinations of *e*_1_ and *e*_*2*_ for a 90% efficacy in reducing symptomatic cases. Because of lack of empirical data, we assume that vaccine efficacy is equally attributable to infection and disease reduction in the main projections. The sterilising immunity after vaccination has been demonstrated. For example, an observational study in the UK (SIREN) found that the risk of being infected was reduced by 70% in health workers after one dose of the Pfizer-BioNTech vaccine.^21 22^

Vaccination of prioritised individuals began from 8 December 2020 in the UK and around 2 million individuals were vaccinated (mostly with a single dose of Pfizer vaccine) by 10 January 2021.^23^ For simplicity, we assume that the mass vaccination starts from 1 January 2021 with a 80% coverage of eligible individuals. We assume that the maximum number of individuals vaccinated per day is 300,000 in England, which matched well with the actual number of vaccinated individuals (mostly with a single dose) in England according to the official statistics. The mass vaccination is modelled as an age-based phase approach, starting from people aged ≥ 70, followed by individuals aged 60-69, 50-59, 20-49, and then those aged 16-19. Although both Pfizer-BioNTec and AstraZeneca vaccines are 2-dose regimens, the policy in the UK has been to initially provide the first dose to as many individuals as possible to maximise the public health impact. Exploratory assessment of data from clinical trials found that the short-term vaccine efficacy from the first dose of the Pfizer-BioNTech vaccine and the AstraZeneca vaccine is, respectively, about 90% and 70%.^24^ For simplicity, we did not separate single or double dose vaccination, and assume that the overall vaccine efficacy is 70% or 90%, and the protection effects start 14 days after vaccination.

Available evidence has indicated that the duration of sterilising (infection protection) immunity after coronavirus infection ranges from 0.5 to two years.^3^ Serum neutralizing antibodies were detected in all participants at four months follow up after SAR-CoV-2 mRNA vaccination.^25^ Therefore, we assume that naturally acquired sterilising immunity lasts for 365 or 730 days, and vaccine-induced sterilising immunity lasts for 182, 365 or 730 days. After waning of sterilising immunity, individuals may be susceptible again to infection with SARS-CoV-2 virus, but the disease reduction immunity is likely longer lasting.^3^ Due to the existence of disease reduction immunity, the reinfectivity of individuals who are reinfected after waning of sterilising immunity may be reduced. Lavine and colleagues estimated that the secondary transmissibility was 0.35 of the primary transmissibility (i.e., the reinfectivity was reduced by 65%).^3^ Evidence from clinical trials and vaccination in the real world indicated that the viral loads and the duration of virus shedding in the infected individuals after vaccination were considerably reduced, compared with unvaccinated individuals.^26 27^ Therefore, we assume that reinfectivity after waning of sterilising immunity is reduced by 20%, 40% or 60%. We also assume that the infectivity of ineffectively vaccinated individuals is the same as recovered individuals after the waning of sterilising immunity. In addition, we assume that vaccination of individuals recovered from natural infection boosts their naturally acquired immunity, which prevents or delays the waning of their sterilising immunity.

In this study, we focus on deaths in people infected with COVID-19, although our model also produces estimates of changes in effective reproduction values (Rt), numbers of infected and vaccinated individuals, and hospitalised patients. We performed multiple simulations under various scenarios. For clarity, we focus on results of selected scenarios in the main text, but report more data on simulation results in Supplementary Tables.

### 4. Model’s mathematical equations

#### Notations

- subscript used: “s” refers to sex, 1: male, 2: female, 3: both male and female; “a” refers to age group, 1: 0-4 years, 2: 5-9 years, 3: 10-19, …, 10: ≥ 80; 11: all age groups. “t” refers to time (day).
- *N*: The number of the population
- λ_s_: Force of infection (*λ*) measures the risk (probability) of infection transmission, which is a function of transmission rate (β) and the prevalence of infectious individuals (*I*) among the population (*N*): *λ=β·I/N*.^1^
- β: The transmission rate β in this discrete-time model is defined as the average number of individuals infected daily by an infectious person. It is a function of the number of daily close contacts per person (c), and the transmission risk per contact between a susceptible and an infectious individual (η): i.e., *β=c·η*. ^2^
- α1: rate of progressing from being exposed to being infectious.
- α2: rate of progressing from being asymptomatic infectious to symptomatic.
- *μ*: proportion of infected individuals who will be symptomatic; age-specific
- *infA*: The fraction of infection force for infected individuals with no or mild symptoms. It was assumed that infA=0.5 in this study.
- fS0: fraction of symptomatic patients who will not be quarantined.
- fSq: fraction of symptomatic patients who will be quarantined (self-isolation).
- fSh: fraction of symptomatic patients who will be hospitalised (including ICU admission).
- γA0: rate of recovering for asymptomatic individuals
- γS0 rate of recovering for symptomatic, not isolated/hospitalised patients
- γS1: rate of being isolated in symptomatic patients
- γS2: rate of recovering in isolated patients
- γH1: rate of being hospitalised for symptomatic patients γ*ϒH2*: rate of recovering in hospitalised patients
- ν_1_: rate of vaccinating susceptible individuals
- ν_2_: rate of vaccinating recovered individuals
- ρ: Relative infectivity of the secondary infection (*I2*) compared with the primary infection (*I1*). For example, if ρ=0.6, the infectivity of *I2* is 40% lower than the infectivity of *I1*
- *e*_1_: Relative efficacy of vaccine for sterilising immunity, reducing risk of virus transmission
- *e*_*2*_: Relative efficacy of vaccine for pathology reduction, reducing the proportion of symptomatic cases after being infected
- IA: Asymptomatic individuals
- IS: Symptomatic patients
- *γ*_*a*_ : Average rate of recovering of asymptomatic individuals
- *γ*_*s*_ : Average rate of recovering of asymptomatic individuals
- *ⴍ*_v_: rate of immunity waning in vaccinated individuals
- *ⴍ*_r_: rate of immunity waning in recovered individuals
- drOth_s,a,t_: sex, age-specific risk of deaths from causes other than covid-19, specific by week of the year.
- drCov_s,a,d_: death risk from infected individuals before recovery, specific according to days since being infected.
- ds0, dsq, and dhos are the proportion of covid-19 deaths among symptomatic patients who are not quarantined, those who are isolated, or hospitalised, respectively. 1=ds0+dsq+dhos

Sex and age specific population:

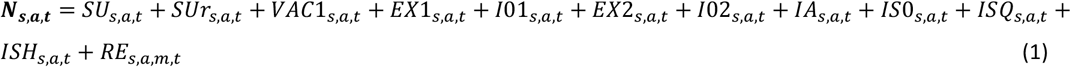

Total number of the primary infection with no symptoms (age-specific):

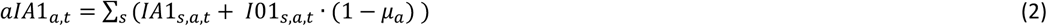

Total number of the primary infections with symptoms, isolated (age-specific):

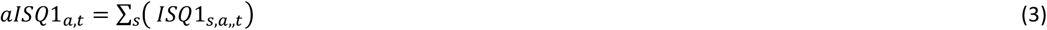

Total number of the primary infections with symptoms, not isolated (age-specific):

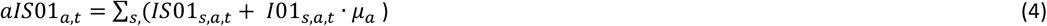

Total number of the secondary infection with no symptoms (age-specific):

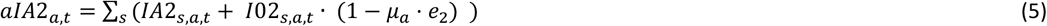

Total number of the secondary infections with symptoms, isolated (age-specific):

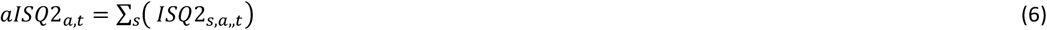

Total number of the secondary infections with symptoms, not isolated (age-specific):

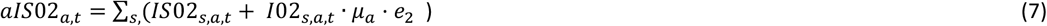

Sex and age specific susceptible population:

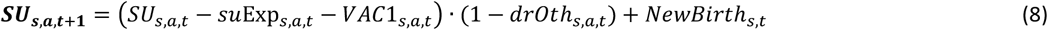

Note: *drOth*_*s,a*,,*t*_ is sex, age-specific death rates for non-covid causes, specific by week of the year.

Newly exposed/infected with SARS-CoV-2 in susceptible individuals:

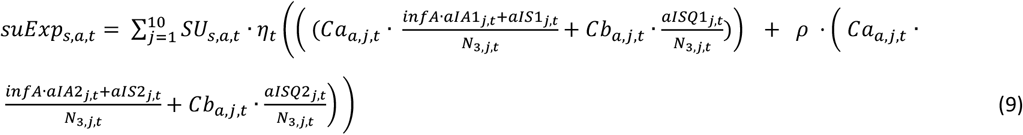

Notes: Ca_a,j,t_ is the average number of general contacts between people aged a and j; and Cb_a,j,t_ is the average number of household contacts between people age a and j.

Newly exposed/infected in vaccinated individuals:

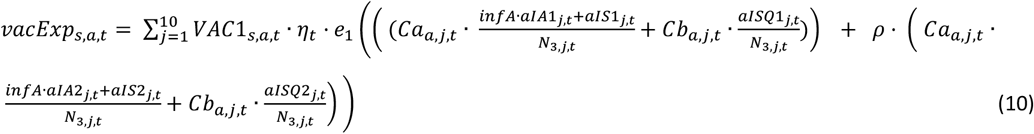

Newly exposed/infected in the recovered or vaccinated after waning of immunity:

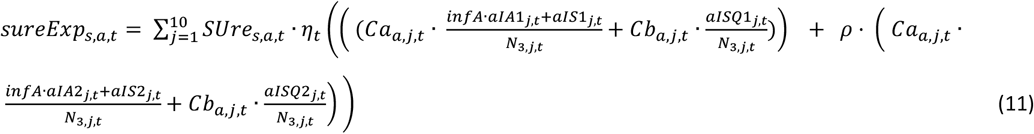

The number of the recovered or vaccinated who lose sterilising immunity (d from 1 to tt):

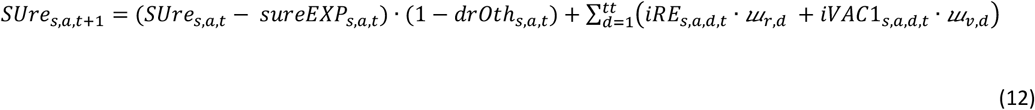

Notes: “*tt*” is the total number of days simulated. *ⴍ*_*r,d*_ and *ⴍ*_*v,d*_ are gamma distributed rate of immunity waning, respectively, a function of days since the recovery and vaccination. *iRE*_*s,a,d,t*_ is the number of recovered since d days from recovery; and *eVAC1*_s,a,d,t_ is the number of vaccinated since d days after vaccination.

The number of new (d=1) primary infections in susceptible individuals:

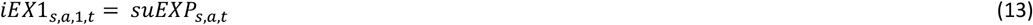

The number of new (d=1) secondary infections in recovered or vaccinated individuals:

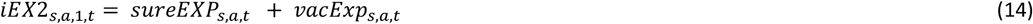

For d=1,2,3…60 (assuming all will be dead or recovered by day 60 after being infected):

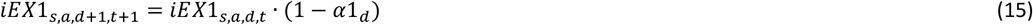

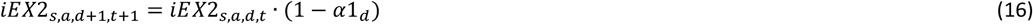

Assumed that covid-19 deaths were from symptomatic patients only. Overall deaths from covid-19 were calculated using the case fatality rates, and timing of covid-19 related deaths were assumed to have a gamma distribution according to days since being infected. Therefore a variable was introduced to record number of symptomatic individuals by days since being exposed/infected to calculate number of covid-19 deaths:

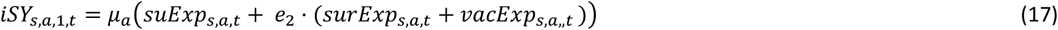

For d=1,2,3…60 (the transmission completed by day 60):

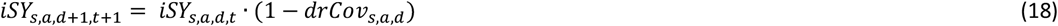

The number of covid-19 deaths at time t:

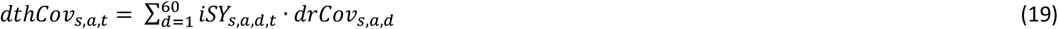

The number of new (d=1) primary infections individuals before onset of symptoms:

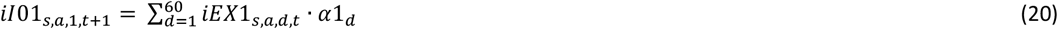

The number of new (d=1) secondary infectious individuals before onset of symptoms:

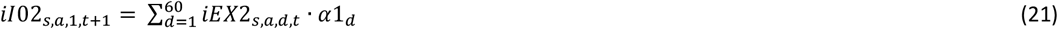

For d=1,2,3…60 (the transmission completed by day 60):

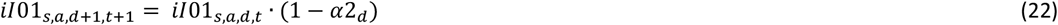

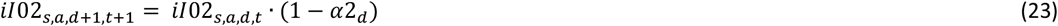

The number of all infectious individuals before onset of symptoms:

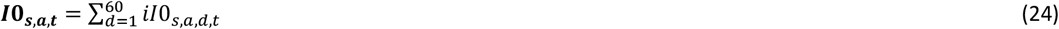

The number of new (d=1) infected individuals with no or very mild symptoms:

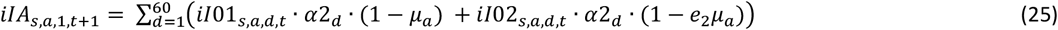

For d=1,2,3…60 (the transmission completed by day 60):

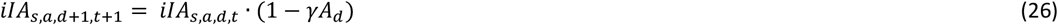

The number of all infectious individuals with no or mild symptoms:

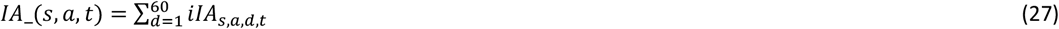

The number of all new (d=1) symptomatic patients:

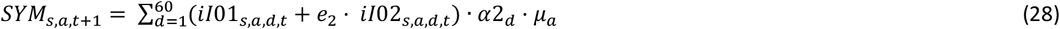

The number of new (d=1) symptomatic patients who are not self-isolated:

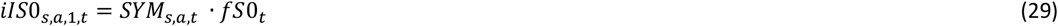

Symptomatic patients (before being isolated or hospitalised:

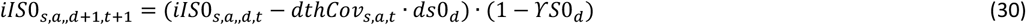

The number of new (d=1) symptomatic patients being isolated/quarantined:

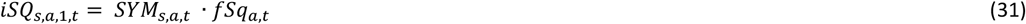

Isolated symptomatic patients:

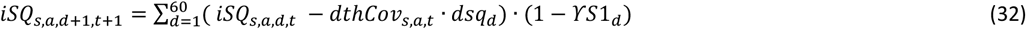

The number of new (d=1) symptomatic patients being hospitalised:

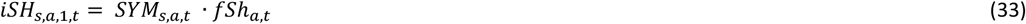

Hospitalised symptomatic patients:

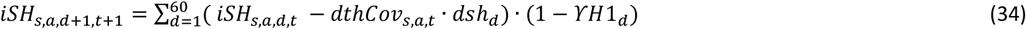

The number newly recovered people (d=1):

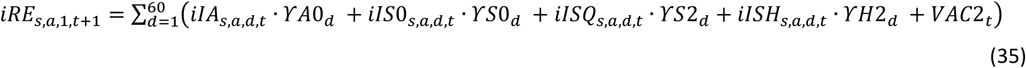

Note: *VAC2*_*t*_ is the number of newly vaccinated individuals who recovered from previous infections.

All recovered for d=1,2,3…tt:

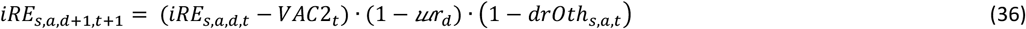

All recovered individuals:

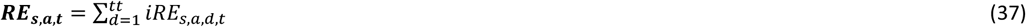

#### Derived reproduction values (R0, Rt)

The basic reproduction ratio (R0) is defined as the average number of individuals infected by a typical infectious individual in a total susceptible population, and effective reproduction ratio (Rt) is the number of individuals infected by an infectious individual when only a proportion of the population are susceptible and the disease transmission dynamic may be influenced by control measures.^28^ R values depend on the risk of infection per contact between an infectious and susceptible person, person-to-person contacts between individuals, the rate of transition from exposed to infectious, infectious period, and the prevalence of susceptible individuals in the population.^1^ In this study, we don’t use the reproduction ratio directly in simulating the spread of SARS-CoV-2 virus. To facilitate the understanding of effects of different intervention strategies, we estimated R0 and Rt during the simulation period, based on average values of relevant parameters and the calculation method used in a modelling study by Giordano et al.^19^

Average values of relevant parameters for estimating R values:

Weighted average fraction of symptomatic individuals in all infected individuals:

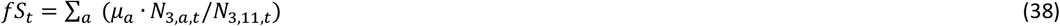

Weighted average fraction of hospitalised symptomatic patients:

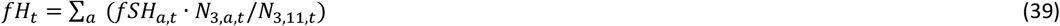

Weighted average fraction of symptomatic patients self-isolated:

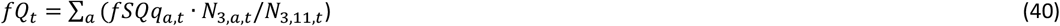

Risk of daily transmission per infectious individual, depending on asymptomatic or symptomatic, household isolated or not:

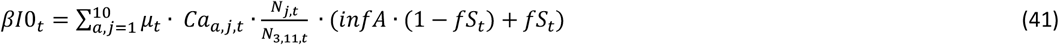

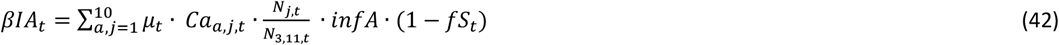

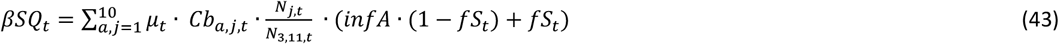

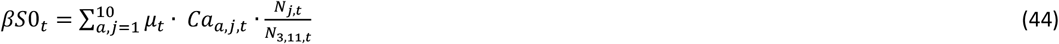

The following transition variables are calculated for estimating R values:

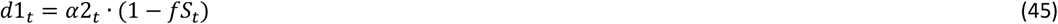

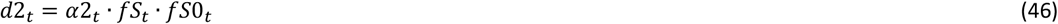

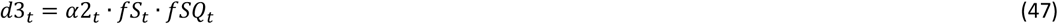

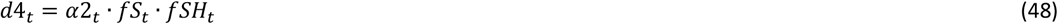

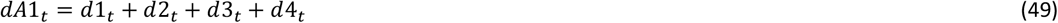

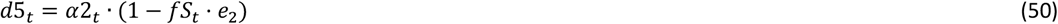

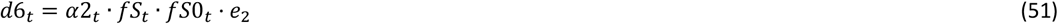

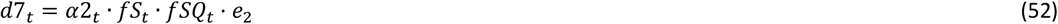

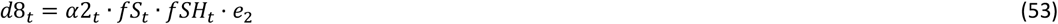

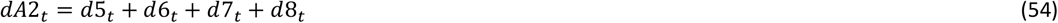

Effective reproductive value (Rt):

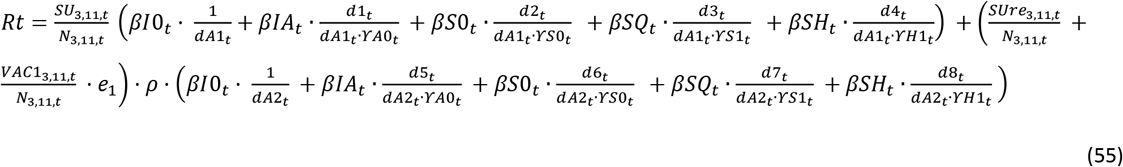

